# Quality and transcript-grounded clinical-risk error burden of AI-generated vs allied health professional clinical notes: a paired simulation study across four disciplines

**DOI:** 10.64898/2026.10.16.26365719

**Authors:** Henry Bergman, Vivian Liu, Sarah Ali

## Abstract

**Background:** Ambient artificial-intelligence (AI) scribes are being deployed beyond medicine into allied-health practice, but comparative evidence mainly concerns physician documentation.

**Objective:** To compare quality and transcript-grounded clinical-risk error burden of AI-generated and allied health professional notes from identical simulated consultations.

**Methods:** Paired simulation across dietetics, physiotherapy, occupational therapy and speech and language therapy (five consultations each; 20 pairs; 40 notes). Heidi generated an unedited first-draft note and a professional documented each encounter. Professionals rated PDQI-9 quality. Professional and automated review identified unsupported statements and omissions; two language models graded clinical risk. Co-primary outcomes were PDQI-9 score and Critical+High error density.

**Results:** Mean PDQI-9 scores were 38.45 and 35.05 for AI and professional notes (paired difference +3.40, 95% CI +0.01 to +6.79; Wilcoxon p=0.042; dz=0.47). The Bayesian estimate was +3.47 (94% equal-tailed credible interval +0.66 to +6.30; P[AI>professional]=0.989). Automated-reviewer all-error density was 3.35 vs 8.55 per note (professional-minus-AI difference 5.20, 95% CI 3.85 to 6.55); omission density was 1.45 vs 5.20 (difference 3.75, 2.80 to 4.65). Critical+High density was 0.35 vs 0.75 (difference 0.40, −0.10 to 1.00). The pooled quality result was driven principally by dietetics.

**Conclusions:** AI-generated notes achieved modestly higher pooled quality and fewer transcript-grounded errors, chiefly omissions; serious-error evidence was inconclusive. Small discipline samples and incomplete ground truth preclude safety or discipline-wide superiority claims.

## Introduction

Ambient artificial intelligence (AI) scribes convert consultation audio into structured clinical documentation. Implementation studies have primarily focused on physician workflow, satisfaction and documentation burden, with generally favourable efficiency but variable output quality and safety assessment.^1–3^ Randomised and prospective evidence suggests that these systems can reduce documentation burden in some settings, although performance and adoption vary across products and clinical contexts.^1–3^ Peer-reviewed commentary have cautioned that deployment has outpaced validation and independent oversight, while the clinical consequences of using ambient scribes rather than other documentation methods remain uncertain.^4,5^

Evidence in allied health is more limited. A 2026 mixed-methods study in DIGITAL HEALTH found high perceived usefulness and ease of use among hospital allied health clinicians, alongside concerns about accuracy, privacy and the need for clinician review.^6^ Commentary has similarly suggested that evidence of adoption is developing more quickly than evidence about the quality of clinical outputs.^7^ This distinction is important because allied health records inform multidisciplinary care and discharge decisions,^8^ while outpatient rehabilitation clinicians experience substantial electronic documentation burden.^9^

Current clinical-note evaluation often combines human quality ratings with counts of unsupported statements, omissions or hallucinations. A recent systematic review found considerable variation in evaluation criteria and widespread use of lexical-overlap measures that were insufficient as standalone indicators of clinical quality.^10^ Human documentation also provides an imperfect comparator. In 217 dictated clinical documents, speech-recognition output contained 7.4 errors per 100 words, falling to 0.4% after transcriptionist review and 0.3% in signed notes. Of these errors, 5.7% in raw output and 6.4% in signed notes were judged clinically significant.^11^

An error taxonomy and clinical-safety framework applied to 12,999 clinician-annotated sentences from large-language-model note generation reported hallucination and omission rates of 1.47% and 3.45%, respectively; major errors fell below previously reported human note-taking rates after prompt and workflow refinement.^12^ In a prospective pilot of 356 physician-reviewed ambient notes, accidental omissions were the most frequent error type (18%), followed by hallucinations (11.5%) and accidental inclusions (9.3%); 5.3% of notes contained an error rated as posing serious or imminent risk.^13^ Comparative quality findings are mixed. One validated outpatient study found marginally lower quality and more hallucinations in ambient notes than in physician reference notes,^14^ while adapted language models have performed strongly in constrained clinical summarisation tasks.^15^ Differences in comparator construction, ground truth and detection sensitivity can therefore materially affect measured performance.

Allied health introduces an additional measurement challenge. Dietetic documentation is often based largely on spoken history and counselling, whereas physiotherapy, occupational therapy, and speech and language therapy also rely on observed, palpated or measured findings that may never enter the consultation audio. A transcript-only reference may therefore classify valid professional observations as unsupported and cannot provide an audio-based scribe with unspoken examination findings. This differential construct coverage is distinct from transcription error. The relevant comparator is the note the scribe would otherwise replace, assessed within the same consultation because encounters vary in complexity and in how much clinical information is spoken aloud.

We therefore conducted a paired simulation in which an AI scribe and a qualified allied health professional documented the same consultation. The co-primary questions were whether note authorship was associated with PDQI-9 quality and whether AI-generated and professional notes differed in Critical+High transcript-grounded error density. Secondary analyses examined variation by discipline, omission and unsupported-statement burden, detection sensitivity, and severity-panel reliability. Estimates apply to the evaluated system configuration and simulated conditions. To our knowledge, no previous paired study has compared AI-generated and allied health professional notes using severity-graded error analysis. Relevant DECIDE-AI items are reported as an adapted framework for this simulated early-stage evaluation; the study did not involve live clinical deployment or human-factors evaluation.^[16]^.

## Methods

### Study design and unit of analysis

We conducted a prospective paired simulation study in the UK across four allied-health disciplines: dietetics, physiotherapy, occupational therapy, and speech and language therapy. Each discipline (“pod”) contributed to five simulated consultations, yielding 20 paired consultations and 40 notes. One AI-generated and one professional-written note were produced independently from each encounter.

### Consultation scripts and simulation procedures

Pairs of independent healthcare professionals served as actors in controlled, discipline-specific scripted consultations. Scripts were synthetic and thematically informed by allied health encounter topics and rewritten to remove identifying information. Scripts incorporated naturalistic complexity, including dysfluency, patient-led tangents and hedging. Consultations were recorded under clean acoustic conditions. The transcript served as the reference standard for note-content assessment but provided incomplete ground truth for examination findings observed or measured without being verbalised.

### Participants, note production and evaluation

Each discipline included one actor pair and two qualified professionals. The professionals alternated between writing and evaluating notes and assessed consultations for which they had not written the professional note. They viewed both notes from the same encounter. Notes carried no explicit AI or professional label, although stylistic differences may have revealed likely authorship. In the dietetics arm, interface text that could reveal authorship was removed before any rating was submitted. Each note received one primary quality rating, with no duplicate-rating substudy. Before the session, evaluators completed an approximately 10-minute training video on the PDQI-9 and adjudication process, and note writers completed a separate approximately 10-minute training video.

### AI system and note production

The AI note was the unedited first draft generated by Heidi for subsequent review by the treating professional. Professionals wrote their notes independently during or immediately after the simulated encounter without later editing of the paired AI note. The study therefore evaluated model output before clinician review rather than the final note ordinarily filed in practice.

### Note-quality outcome

Note quality was assessed using the nine-item Physician Documentation Quality Instrument (PDQI-9). Its items evaluate whether documentation is up to date, accurate, thorough, useful, organised, comprehensible, succinct, synthesised and internally consistent. Each item is scored from 1 to 5, producing a total score from 9 to 45.^17^

### Error identification

Documentation errors comprised unsupported statements, defined as note content not supported by the consultation transcript, and omissions, defined as clinically relevant spoken content absent from the note. Two methods with different expected sensitivities identified errors. In the professional-adjudication arm, evaluators recorded free-text flags that were separated into atomic claims and checked against the transcript; false-positive flags were removed. In the automated-reviewer arm, a large language model compared each transcript and note in both directions before false-positive screening. Free-text responses stating that no issue was identified were normalised deterministically without a model call.

Evaluation stages were blinded to note authorship. The study-specific rubric, configurations and decision rules are reported in Supplementary Methods; verbatim proprietary prompts are not publicly released.

### Clinical-risk grading and the rater panel

Each error was graded using a 1-5 severity by 1-5 likelihood matrix, consistent with the ISO 14971 definition of risk as a combination of harm severity and probability.^18^ Products were classified as Critical (16-25), High (10-15), Moderate (5-9) or Low (1-4). The co-primary safety outcome defined clinically significant errors as Critical+High.

The initial panel comprised GPT-5.5, Claude Opus 4.8 and Gemini 2.5 Pro to reduce reliance on any single model family. Gemini classified 59.2% of 238 errors as Critical+High, compared with 8.4% for GPT-5.5 and 4.2% for Claude. It matched the three-model consensus tier for 34.9% of errors, compared with 80.3% and 84.5%, respectively, and was therefore outvoted for most disagreements.

The declared analysis used the two models that determined the consensus result, GPT-5.5 and Claude Opus 4.8. When they disagreed, the harsher tier was retained because a two-rater panel had no majority. This conservative rule was applied equally to both note types. Full three-model marginal results are reported in Supplementary Table S5. Because the panel was changed after reviewing model outputs, this decision was post hoc. Reliability was summarised using exact four-tier agreement, observed agreement for severe vs non-severe classification, and Cohen’s kappa for the binary classification. Four-tier kappa was calculated only for the declared two-rater panel. Given the rarity of severe errors, kappa was interpreted alongside observed agreement because of its sensitivity to imbalanced marginals.^19,20^ Four-tier kappa fell within the range conventionally described as fair,^21^ while severe vs non-severe agreement exceeded 90% in both detection arms. Models received no explicit authorship label, although stylistic cues may have remained. GPT-5.5 was used both in the automated detection pipeline and in the severity panel, so those stages were not statistically or methodologically independent.

### Outcomes

Co-primary outcomes were PDQI-9 total score and Critical+High errors per note under each detection method. Severe-error burden was expressed as error density because, with 20 notes per group, a binary indicator of any severe error would be strongly influenced by whether several errors occurred within one note. Density retained the complete error count and denominator. Per-note counts are provided in the Supplementary Materials to support alternative summaries. Secondary outcomes included PDQI-9 item scores, dispersion, the proportion of scores below 32, discipline-specific quality, total-error density, unsupported-statement and omission density, mean severity per error, detection sensitivity and precision, false-positive rates, and severity-panel reliability.

Detailed item-level, sensitivity, subgroup, error-register, calibration, model-diagnostic and visual results are reported in Supplementary Tables S1-S24 and Supplementary Figure S1.

### Statistical analysis

Analyses followed the reporting framework specified for the companion multi-country study, adapted to the allied health design.^22^ The primary quality comparison used a paired Wilcoxon signed-rank test on consultation-level PDQI-9 totals. The mean paired difference was accompanied by a t-based 95% confidence interval and Cohen’s dz; a paired t test was also reported. Item-level paired Wilcoxon tests used Benjamini-Hochberg false-discovery-rate correction, and dispersion was compared using Levene’s test. No multiplicity adjustment was applied across the two co-primary outcomes; results are therefore interpreted through effect estimates and uncertainty rather than a joint confirmatory decision. An exploratory distribution-based threshold of 0.5 x pooled SD was used because no validated PDQI-9 minimal important difference is available; it is not an anchor-based estimate for this measure or population.^23^

The Bayesian model was PDQI-9 total ∼ note type + (1|consultation) + (1|evaluator) + (1|note writer), with a Gaussian likelihood and identity link. Each AI note carried the identifier of the professional who wrote its paired comparator; for AI notes, the note-writer term therefore represented clustering by the paired professional rather than AI writing skill. The primary fit used bambi 0.18.0 with PyMC 6.0.1, two chains, 1,000 post-warm-up draws per chain, 1,000 tuning steps, target acceptance 0.90 and seed 20260518. Default weakly informative, data-scaled priors were used and were not tuned to this study. The note-type coefficient is reported with the analysis pipeline’s 94% equal-tailed credible interval (3rd to 97th posterior percentiles), not a highest-density interval. No imputation was required: one empty incomplete record was excluded, leaving 40 complete notes and 20 complete pairs.

Convergence, posterior predictive checks, assumptions and alternative specifications are reported in Supplementary Tables S17-S20.

Sample size was determined by the feasibility of recruiting two qualified professionals and one actor pair per discipline and completing five scripted consultations per discipline while preserving the cross-evaluation design. With 20 pairs, the minimum detectable effect at 80% power for a two-sided paired t test at alpha=0.05 was dz=0.66, equivalent to 4.8 PDQI-9 points at the observed paired-difference SD of 7.2. This exceeded the observed dz=0.47; inferential results are therefore exploratory. Planning values are reported in Supplementary Table S22.

For error-density contrasts, 95% confidence intervals were estimated using a paired bootstrap that resampled consultations 10,000 times. The primary safety contrast is reported as both the professional-minus-AI difference and the professional-to-AI ratio. Per-note severe-error counts were also compared using a paired Wilcoxon signed-rank test. These analyses are reported in Supplementary Table S23.

With five consultations per discipline, the smallest possible two-sided Wilcoxon signed-rank p value is 0.0625. Within-discipline tests therefore could not reach alpha=0.05, regardless of effect magnitude or consistency. The four discipline-specific p values were adjusted using the Benjamini-Hochberg procedure. Discipline-specific results are presented as the number of pairs favouring each note type, mean paired differences, confidence intervals and point estimates of effect size. Percentile bootstrap intervals for Cohen’s dz were not reported because resamples with near-zero paired standard deviations produced degenerate estimates. All discipline-specific inferential analyses were exploratory.

Frequentist analyses were conducted in Python using pandas and scipy; Bayesian analyses used bambi 0.18.0, PyMC 6.0.1 and ArviZ 1.1.0. The automated reviewer and severity panel used a pinned, version-controlled pipeline and a frozen data snapshot. Each stage generated a separate output, and severity scoring required a grade from every declared panel member. Analysis provenance is summarised in Supplementary Tables S16 and S24.

### Ethics and consent

All participants, including actors, note-writing allied health professionals and evaluators, provided written informed consent to participate and for the research use of the resulting notes and recordings. No real patients participated, and no patient-identifiable data were used. Scripts were thematically informed by real clinician encounter topics but were rewritten without original identifying information, making them synthetic and non-reidentifiable to the source encounters. The UK Health Research Authority decision tool indicated that NHS Research Ethics Committee review was not required for the UK site; it does not constitute approval or a waiver by an institutional research ethics committee.^24^ No patient or public involvement contributed to the design, conduct, reporting, interpretation or dissemination of this study.

## Results

### Note quality: co-primary outcome

AI-generated notes had a mean PDQI-9 score of 38.45, compared with 35.05 for professional notes. The mean paired difference was +3.40 points (95% CI +0.01 to +6.79), with Wilcoxon p=0.042, paired t p=0.049 and Cohen’s dz=0.47. The Bayesian crossed-random-effects model estimated a +3.47-point note-type coefficient (94% equal-tailed credible interval +0.66 to +6.30), with P(AI>professional)=0.989. The estimate was close to the unadjusted paired difference.

The pooled quality difference varied by discipline (Table 1 and Figure 1). Mean paired differences were +11.0 points in dietetics, +2.8 in physiotherapy, +1.0 in occupational therapy and −1.2 in speech and language therapy. Thirteen of 20 paired comparisons favoured AI-generated notes. Excluding dietetics reduced the pooled difference to +0.87 (95% CI −2.35 to +4.08; p=0.489), while excluding the two largest paired differences reduced it to +2.00 (95% CI −0.93 to +4.93; p=0.129) (Supplementary Table S2). In 45% of pairs, the AI score exceeded the professional score by at least the exploratory 3.65-point distribution-based threshold; this is not a validated minimally important difference for PDQI-9.

**Figure 1.**
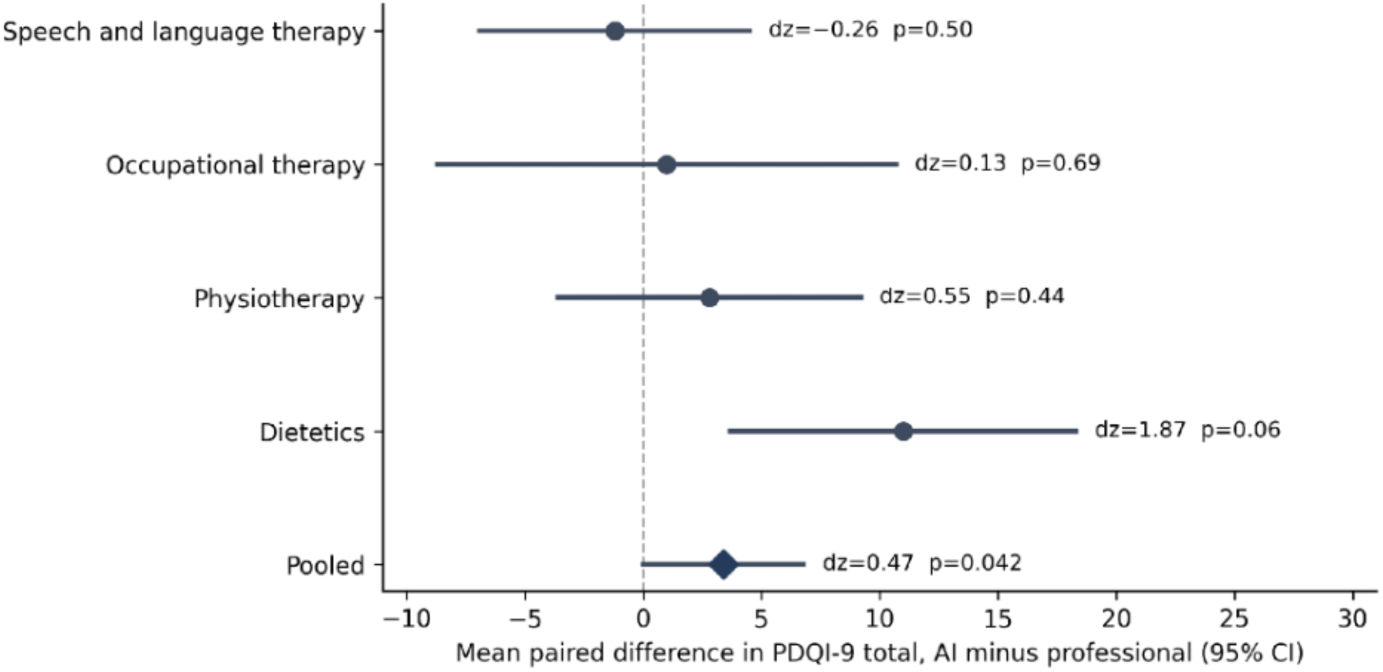
Mean paired PDQI-9 difference with t-based 95% confidence interval by discipline and pooled. Positive values favour AI. Discipline-level p values shown in the figure are raw; Table 1 reports Benjamini-Hochberg-adjusted p values. At n=5, the two-sided Wilcoxon floor is 0.0625.

**Table 1.** PDQI-9 quality by discipline (n=5 consultations per discipline).

| Discipline | AI | Prof. | $\Delta$ | 95% CI | $d_z$ | Pairs favouring AI | p (BH-adj) |
| --- | --- | --- | --- | --- | --- | --- | --- |
| Dietetics | 42 | 31 | 11 | +3.7 to +18.3 | 1.87 | 5/5 | 0.25 (ns) |
| Physiotherapy | 43.6 | 40.8 | 2.8 | -3.6 to +9.2 | 0.55 | 3/5 | 0.67 (ns) |
| Occupational Therapy | 29.2 | 28.2 | 1 | -8.7 to +10.7 | 0.13 | 3/5 | 0.69 (ns) |
| Speech and Language Therapy | 39 | 40.2 | -1.2 | -6.9 to +4.5 | -0.26 | 2/5 | 0.67 (ns) |
| Pooled | 38.45 | 35.05 | 3.4 | +0.01 to +6.79 | 0.47 | 13/20 | 0.042 |

The model intercept was 38.1. The posterior SD was 1.12 for consultation, and the combined professional-level SD for evaluator and note writer was 7.4 (94% credible interval 4.1 to 15.7). Evaluators and note writers were fully nested within discipline, and the same two professionals wrote and evaluated notes within each discipline, so their variance components were not separately interpretable and are reported jointly. Professional-level variation exceeded both the note-type coefficient and consultation-level variation and also absorbed between-discipline differences. Adding discipline as a fixed effect did not materially change the note-type estimate (+3.46; Supplementary Tables S20-S21).

For the primary fit, R-hat was at most 1.01 and minimum bulk and tail effective sample sizes were 334 and 547. Four of 2,000 post-warm-up transitions diverged (0.2%). Posterior predictive checks reproduced the observed means, dispersion and paired difference; the principal misfit was the bounded PDQI-9 ceiling, with 13.7% of Gaussian-replicated scores exceeding 45. A right-censored sensitivity model estimated an AI advantage of +3.50 points (94% equal-tailed credible interval +0.11 to +6.95). Across prior, likelihood, ceiling and frequentist sensitivity analyses, point estimates ranged from +3.19 to +3.50 (Table 2; Supplementary Tables S18-S20).

**Table 2.** Robustness of the estimated AI advantage in PDQI-9 score. ETI, equal-tailed credible interval; CI, confidence interval; REML, restricted maximum likelihood.

| Specification | AI advantage | Interval | $P(\text{AI} > \text{professional})$ | Divergences |
| --- | --- | --- | --- | --- |
| Primary Bayesian model | +3.47 | 94% ETI +0.66 to +6.30 | 0.989 | 4 |
| Diagnostic refit | +3.44 | 94% ETI +0.65 to +6.18 | 0.988 | 10 |
| Wide priors | +3.45 | 94% ETI +0.69 to +6.28 | 0.989 | 1 |
| Regularising priors | +3.19 | 94% ETI +0.49 to +5.86 | 0.986 | 0 |
| Student-t likelihood | +3.43 | 94% ETI +0.54 to +6.32 | 0.987 | 0 |
| Right-censored at 45 | +3.50 | 94% ETI +0.11 to +6.95 | 0.973 | 4 |
| Discipline fixed effect | +3.46 | 94% ETI +0.56 to +6.32 | 0.987 | 19 |
| Consultation random effect only | +3.39 | 94% ETI +0.08 to +6.80 | 0.973 | 0 |
| Frequentist REML, crossed components | +3.46 | 95% CI +0.83 to +6.09 | p=0.010 | Not applicable |

PDQI-9 dispersion was similar between groups (SD 7.21 for AI-generated notes and 7.19 for professional notes; Levene $p=0.46$ ).

None of the nine PDQI-9 item comparisons remained statistically significant after false-discovery-rate correction; the smallest adjusted p value was 0.060. Unadjusted mean differences favoured AI-generated notes for every item and ranged from +0.20 to +0.60 (Supplementary Table S1).

### Transcript-grounded error burden: co-primary outcome

The automated reviewer identified 238 errors across 40 notes. AI-generated notes contained 3.35 errors per note, compared with 8.55 in professional notes. The professional-minus-AI difference was 5.20 errors per note (95% CI 3.85 to 6.55), and the professional-to-AI ratio was 2.55 (95% CI 1.93 to 3.59). The contrast arose mainly from omissions: 1.45 versus 5.20 per note, a difference of 3.75 (95% CI 2.80 to 4.65) and ratio of 3.59 (95% CI 2.47 to 6.06). Critical+High densities were 0.35 and 0.75 per note, respectively; the difference was 0.40 (95% CI −0.10 to 1.00) and the ratio was 2.14 (95% CI 0.67 to 4.60), so the serious-error contrast was imprecise and compatible with no difference. Mean severity per error was 6.90 for AI-generated notes and 7.50 for professional notes. Error composition by note is reported in Supplementary Table S15.

In the automated-reviewer arm, AI-generated notes contained 10 Low, 50 Moderate, 7 High and no Critical errors; professional notes contained 21 Low, 135 Moderate, 14 High and 1 Critical error. The Critical error recorded a left middle cerebral artery stroke as the admission diagnosis when the transcript supported only a general stroke context. The two panel models assigned different tiers, and the declared two-rater rule retained the more severe classification.

Targeted review of the 22 Critical+High automated-reviewer flags classified 16 as genuine, including 6 in AI-generated notes and 10 in professional notes. Confirmed severe-error densities were therefore 0.30 and 0.50 per note, respectively. Three additional flags were classified as uncertain, two as reasonable inference and one as appropriate concision (Supplementary Table S13). At least one severe error occurred in 7 AI notes and 6 professional notes. The professional note contained more severe errors in 6 consultations, the AI note in 4, and 10 were tied (paired Wilcoxon p=0.22), indicating that the higher professional density reflected clustering within a small number of notes. These screened estimates differ from the pre-screen densities reported in Table 3.

**Table 3.**
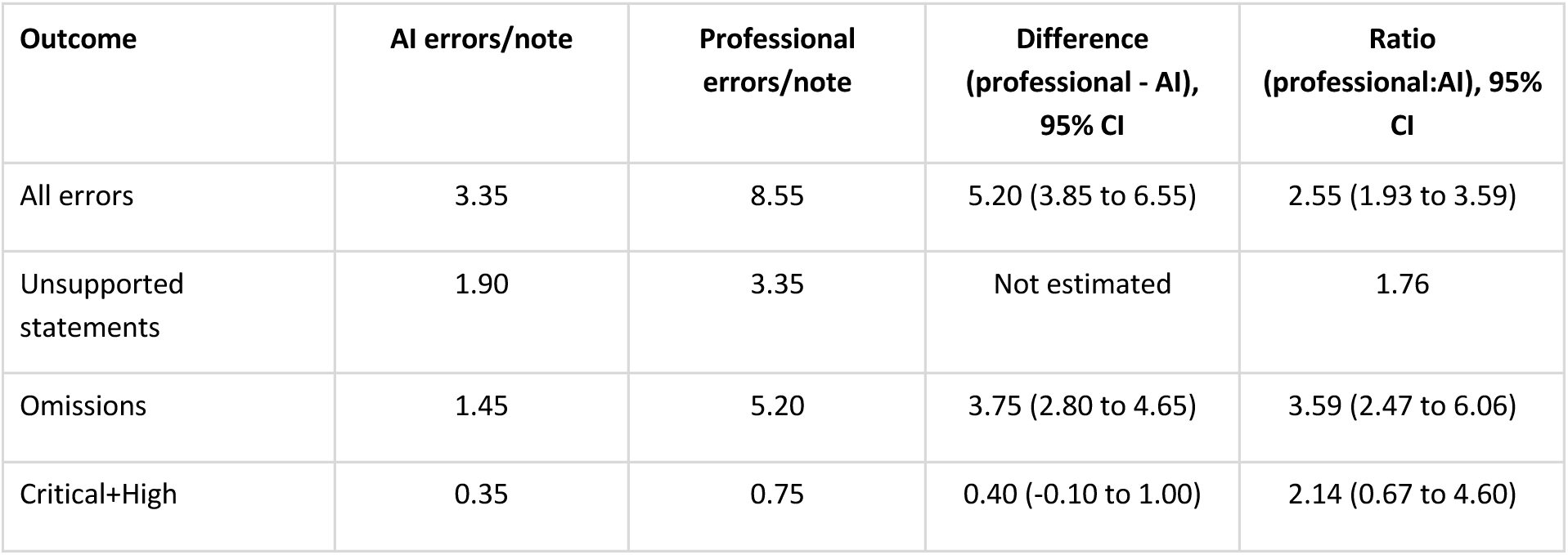
Transcript-grounded error density by note author, automated reviewer (20 notes per group). Confidence intervals were estimated by paired bootstrap resampling of consultations (10,000 resamples).

| Outcome | AI errors/note | Professional errors/note | Difference (professional - AI), 95% CI | Ratio (professional:AI), 95% CI |
| --- | --- | --- | --- | --- |
| All errors | 3.35 | 8.55 | 5.20 (3.85 to 6.55) | 2.55 (1.93 to 3.59) |
| Unsupported statements | 1.90 | 3.35 | Not estimated | 1.76 |
| Omissions | 1.45 | 5.20 | 3.75 (2.80 to 4.65) | 3.59 (2.47 to 6.06) |
| Critical+High | 0.35 | 0.75 | 0.40 (-0.10 to 1.00) | 2.14 (0.67 to 4.60) |

### Error burden by discipline

Critical+High density was lower for AI notes in physiotherapy (0.40 vs 1.20), occupational therapy (0.20 vs 0.40), and speech and language therapy (0.40 vs 1.20), and higher in dietetics (0.40 vs 0.20). The dietetics reversal represented two AI severe errors and one professional severe error. Overall error frequency favoured AI in each discipline, from 1.5x in speech and language therapy to 3.9x in dietetics (Supplementary Table S3 and Figure 2).

**Figure 2.**
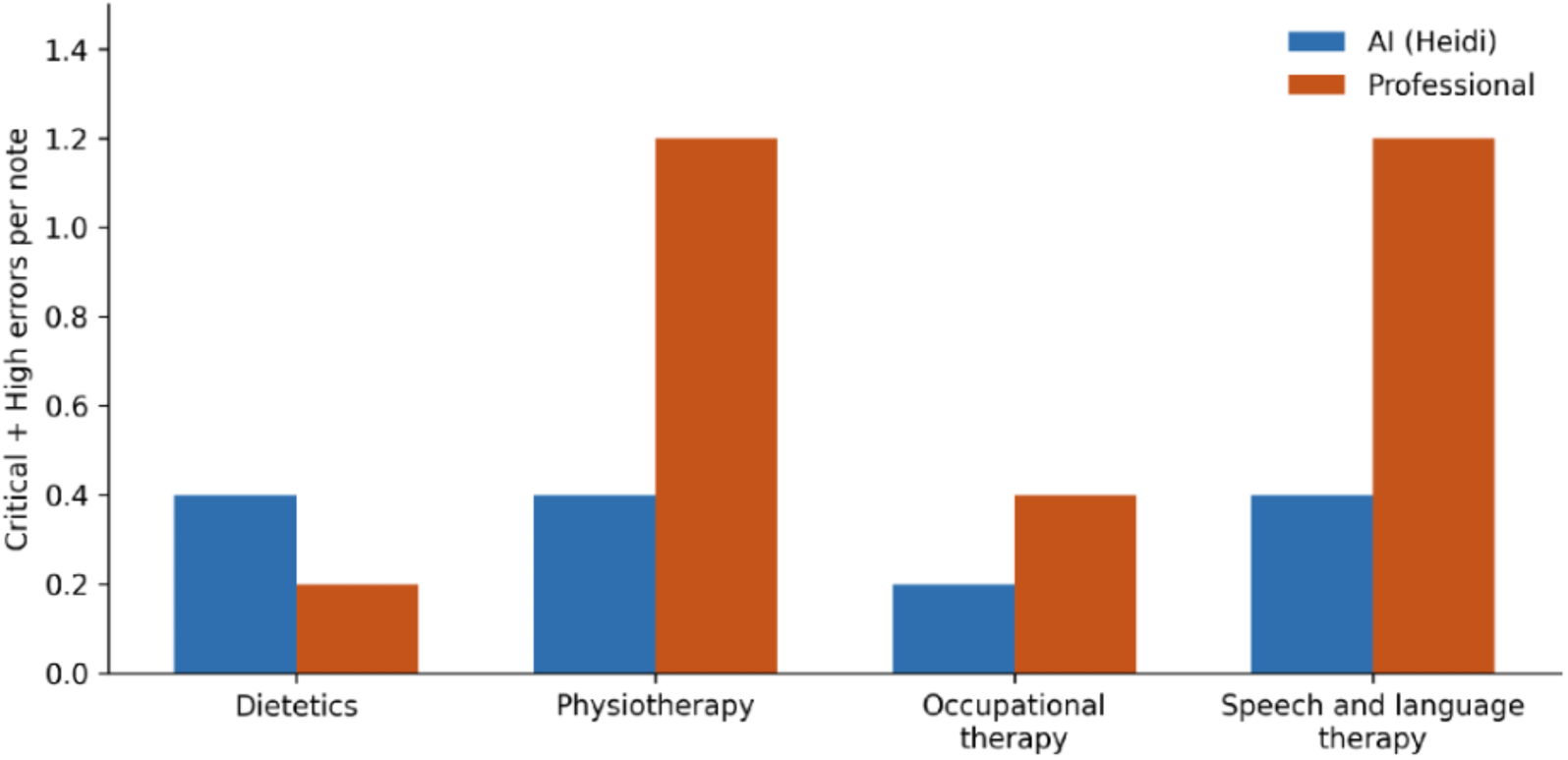
Critical+High error density by discipline and note author, automated reviewer. Each discipline included five notes per group. Discipline-specific estimates are descriptive. For the pooled comparison, the professional-minus-AI difference was 0.40 errors per note (95% CI −0.10 to 1.00) and the professional-to-AI ratio was 2.14 (95% CI 0.67 to 4.60).

### Professional-adjudication arm and detector performance

Professional adjudication confirmed 50 errors. AI-generated notes contained 0.95 errors per note, compared with 1.55 in professional notes. Critical or High error densities were 0.00 and 0.30 per note, comprising no severe errors in AI-generated notes and six severe errors across three professional notes. Mean severity per error was 5.58 and 7.48, respectively. Each detection method identified one Critical error in a professional note.

Using automated-reviewer classifications as the study reference, professionals identified 8.4% of confirmed unsupported statements and 7.4% of confirmed omissions. Precision for professional unsupported-statement flags was 38.5% for AI-generated notes and 88.9% for professional notes. Omission precision was 66.7% and 71.1%, respectively, while omission sensitivity was 10.0% and 6.7%. These measures describe agreement with the study reference procedure rather than diagnostic accuracy against independent clinical ground truth.

The automated reviewer’s false-positive rate for omission flags was 34.5% for AI-generated notes and 19.2% for professional notes; 7 of 29 AI omission flags reflected appropriate concision. False-positive rates for unsupported statements were 7.9% and 1.5%, respectively, while 78.9% and 77.6% were classified as genuine (Supplementary Tables S11 and S12).

### Severity-panel reliability

Exact four-tier agreement between GPT-5.5 and Claude Opus 4.8 was 66.0% in the automated-reviewer arm and 58.0% in the professional-adjudication arm. Agreement for severe versus non-severe classification was 92.9% and 90.0%, with Cohen’s kappa values of 0.339 and 0.260, respectively (Table 4). High observed agreement with lower kappa is compatible with sparse severe-event marginals.^19,20^ These statistics describe agreement between the models and do not establish clinical validity.

**Table 4.** Severity-panel reliability.

| Measure | Calibrated arm (n=238) | Professional arm (n=50) |
| --- | --- | --- |
| Exact four-tier agreement | 66.00% | 58.00% |
| Agreement, severe vs not | 92.90% | 90.00% |
| Cohen's $\kappa$, severe vs not | 0.339 | 0.26 |

## Discussion

In this paired simulation, AI-generated notes achieved modestly higher pooled PDQI-9 scores and lower total transcript-grounded error density than professional notes from the same consultations. The pooled quality difference was sensitive to the inclusion of dietetics. Confidence intervals for the all-error and omission contrasts excluded the null. The Critical+High contrast favoured AI in direction but remained imprecise, and the paired per-note comparison was not statistically significant. Mean severity per detected error was similar between note types. The direction of the descriptive findings was consistent across both detection methods, but neither method provides independent clinical ground truth for every error or severity classification.

### Interpretation and plausible mechanisms

Differences between disciplines are central to interpreting these findings. Dietetics showed the largest quality advantage for AI-generated notes, whereas differences were small or negative in the three examination-focused disciplines. Information availability offers one plausible explanation: most dietetic history and advice are communicated verbally and are therefore accessible to an audio-based system. By contrast, physiotherapy, occupational therapy, and speech and language therapy also generate clinically relevant findings through observation, palpation, measurement and task performance. These findings may be absent from both the scribe input and the transcript used for adjudication.

This mechanism predicts two consequences, both of which are reflected in the data. However, the observed error profile is compatible with the proposed mechanism rather than proof of it. Without a multimodal reference record and independent adjudication by clinicians from each discipline, the extent to which the pattern reflects measurement artefact cannot be determined.

First, AI error rates would be expected to increase with reliance on physical examination because the system receives an incomplete representation of the encounter. AI errors per note increased from 1.60 in dietetics to 2.60 in physiotherapy, 3.40 in occupational therapy and 5.80 in speech and language therapy. Occupational therapy had the highest AI omission rate of any discipline, at 2.80 per note and 3.5 times the dietetics rate, consistent with functional-assessment findings being performed but not verbalised. However, the composition of AI errors did not follow this pattern uniformly.

Speech and language therapy had the highest AI unsupported-statement rate in the study, at 4.80 per note and six times the dietetics rate, despite an unremarkable omission rate. One hypothesis is that a scribe generating a structured note may populate expected assessment and plan sections even when the relevant information was performed rather than spoken, thereby replacing a potential omission with plausible but unsupported content. This remains a hypothesis rather than a finding. The data suggest that examination dependence may increase AI error rates without producing a consistent error-type signature, but five consultations per discipline are insufficient to distinguish this mechanism from discipline-specific or scenario-specific effects.

Second, transcript-grounded assessment would be expected to generate a disproportionate number of flags against professional-written notes in examination-focused disciplines. Human unsupported-statement flags were 2.6 to 3.3 times more frequent in the three examination-focused disciplines than in dietetics, at 3.60 to 4.60 vs 1.40 per note. By contrast, human omission rates were broadly similar across disciplines, at 5.20 to 5.60 vs 4.80 per note, or 1.1 to 1.2 times the dietetics rate (Supplementary Table S7 and Supplementary Figure S1). Examination dependence therefore appeared to increase one type of human-note flag while leaving the other largely unchanged.

Inspection of the flag register identified two possible explanations with different implications. Some flags clearly reflected measurement artefact because clinicians had documented information that was observed, measured or otherwise known but absent from the transcript. Examples included encounter dates and times in physiotherapy notes, documentation of consent, a night-time pain score of 0/10 when the transcript stated only that night pain was “okay”, and paraspinal spasm when the transcript described tightness. Other flags may represent genuine and clinically consequential documentation errors. Several concerned functional status being recorded as independent when the transcript documented assistance, including sitting at the edge of the bed being described as independent immediately after the physiotherapist offered help, and assistance of one being recorded for all activities of daily living when the transcript supported only cueing during assessment. If these statements reflected the clinician’s broader judgement of baseline function, they may be artefacts of the transcript-only reference. If they overstated the patient’s independence, they represent errors with potential consequences for handover and discharge planning.

The flags identified in dietetics were qualitatively different. They primarily involved conflation or misattribution of spoken information, including documenting a phosphate binder when a potassium binder was being considered, conflating four days without eating with four days of intravenous fluids, and attributing a supplement flavour to the patient rather than her husband. These are errors involving information contained in the conversation and are therefore well suited to transcript-grounded detection. Dietetics consequently provides the most interpretable comparison of the four disciplines, despite including only five consultations. Importantly, the discipline driving the pooled quality difference was also the discipline in which the transcript-only design was least likely to disadvantage the AI system.

The wider methodological implication is that a consultation transcript provides incomplete ground truth in examination-focused allied health disciplines, while the consultation audio provides incomplete input to the scribe. Transcript-grounded evaluation may therefore misclassify both note types: AI-generated notes may be penalised for omitting information unavailable in the audio, while professional-written notes may be penalised for including valid observations unavailable in the transcript. The magnitude and direction of these effects cannot be established without independent clinical adjudication. The deployment implication is narrower but clearer: an ambient scribe can capture examination findings when they are verbalised as they are obtained; when they remain unspoken, the system has no source material from which to document them.

Alternative explanations remain plausible. The observed discipline pattern may reflect the selected scenarios, documentation templates, writer behaviour or evaluator severity rather than stable differences between professions. Bayesian variance estimates indicated that writer and evaluator effects were large relative to the note-type coefficient, although both were estimated from few levels and writers were nested within disciplines. Post hoc exclusion of one severity model may also have affected tier allocation. These features support estimation and hypothesis generation but preclude confirmatory claims. False-positive calibration cannot resolve the underlying artefact because the calibrating model relies on the same transcript and therefore shares the same blind spot.

Distinguishing valid clinical observations from unsupported claims requires independent review by clinicians from the relevant disciplines against ground truth that includes examination findings. Such a substudy is feasible for the 60 relevant flags and represents the single most valuable extension of this work.

### Relationship to previous evidence

The study extends allied health research that has so far concentrated on acceptability, workflow and documentation burden.^6–9^ Its paired output comparison addresses a different question: the quality and transcript-grounded error burden of notes created from identical encounters. The pooled direction differs from an outpatient physician study in which ambient notes scored marginally lower in overall quality than physician reference notes on a modified PDQI-9 (4.20 vs 4.25 of 5) and were more often affected by hallucination (31% vs 20% of notes).^14^ This difference may arise from comparator construction, clinical setting, note templates, product configuration and error-detection method. A recent emergency-department comparison similarly found AI notes to be similar or lower in quality than human-scribe notes, with greater physician editing contribution.^25^ Numerical comparisons across studies must therefore account for both the comparator and the measurement instrument.

The detection-sensitivity analysis further demonstrates how strongly reported error rates depend on the assessment method. Allied health professionals identified fewer than 10% of the errors subsequently confirmed by the automated reviewer. Error counts derived from unaided human review are therefore not directly comparable with those produced by high-sensitivity automated review followed by false-positive screening. The companion multi-country study of the same AI system against junior-to-middle-grade clinicians, using the same analytical framework, reported a larger quality difference than the present study, at +5.08 PDQI-9 points (dz=0.55) vs +3.40. This difference may reflect the professional comparator, setting, scenarios or other design factors rather than differential system performance. The companion study is a preprint and has not undergone peer review.^22^

Previous allied health literature has examined whether clinicians will accept ambient documentation and whether it reduces their workload, but has provided limited evidence about the quality of the resulting notes. The present findings suggest quality parity to modest advantage, accompanied by fewer omission-related errors. This represents a more measured value proposition than that supported by the physician comparison and should be communicated accordingly.

The wider evaluation literature supports a layered assessment. A 2026 systematic review found that lexical metrics, human criteria and LLM-based evaluators varied substantially across clinical-note studies and recommended combined semantic, model-based and targeted human review.^10^ Peer-reviewed work on LLM judges has demonstrated high agreement with expert ratings in some clinical summarisation tasks,^26^ while methodological reviews emphasise task-specific failure modes and the need for human oversight.^27^ The present panel achieved high agreement for severe-vs-not classification and lower agreement across four tiers, illustrating the difference between reliability and validity. A reliable panel may still share systematic errors, particularly when all evaluators are grounded on the same incomplete transcript. A proof-of-concept study scoring clinical documentation with three language models on a modified PDQI-9 reported high absolute scores alongside negative inter-rater agreement (Krippendorff’s alpha=-0.433), indicating disagreement sufficient to preclude conclusions about documentation quality from those scores alone.^28^ Automated severity grading in the present study is therefore reported as a reliability statistic and not as a validated substitute for expert clinical adjudication.

### Clinical and methodological implications

The findings support further testing of clinician-reviewed ambient documentation in allied health workflows, evaluated separately by discipline. The defensible interpretation is modestly higher pooled quality and fewer transcript-grounded omissions under the simulated conditions; the data do not establish a reduction in serious clinical errors. Professionals reviewing AI notes raised unsupported-statement flags that often did not agree with the automated study reference and missed most reference flags, so review performance should be described as agreement with that procedure rather than clinical diagnostic accuracy.^4^ In this dataset, AI failures included over-specification of spoken content, such as ‘spicy fruit’ for ‘very spicy food’ and bilateral amputations for two toes. Review protocols could therefore prioritise specific, checkable clinical details, omissions and claims about functional independence. The results concern unedited drafts and simulated risk and do not establish reduced patient harm or the safety of deployed notes. Studies of filed notes should quantify clinician corrections and residual errors after review. Signed documentation is subsequently copied, summarised, coded and reused, so safety evaluation should also examine downstream propagation.^31^ Patient-facing AI-assisted documentation raises related questions about provenance, error contestation and accountability once a machine-generated draft enters a shared record.^32^

Methodologically, transcript-grounded evaluation requires explicit construct boundaries. In speech-led encounters, the transcript may approximate the note’s source material. In examination-led encounters, the reference should include structured observations, measured values and task-performance findings. Multimodal ground truth would permit separation of unsupported statements from valid but unspoken observations and would make error attribution to the scribe, clinician or reference process more defensible.

### Limitations

This study had several limitations. The sample comprised 20 paired consultations, with five per discipline. No formal sample-size calculation was performed; the minimum detectable effect at 80% power was dz=0.66, larger than the observed quality effect, and the Critical+High contrast was imprecise. Discipline-specific estimates were imprecise, and the pooled quality result was sensitive to exclusion of dietetics. Each note received one primary PDQI-9 rating, evaluators viewed both paired notes and may have inferred authorship, and no inter-rater reliability substudy was conducted.

Evaluators and writers were nested within discipline and their separate variance contributions could not be distinguished from each other or from between-discipline differences, although adding discipline as a fixed effect did not alter the note-type estimate. The Bayesian model had a small number of divergent transitions and an unbounded Gaussian likelihood that did not represent the PDQI-9 ceiling; targeted sensitivity analyses gave similar note-type estimates. PDQI-9 was developed for physician documentation, and its construct validity, measurement invariance and clinically important difference have not been established for allied health notes.^17,29,30^ The transcript provided incomplete ground truth for examination-focused consultations, limiting error detection and false-positive screening. Severity grading relied on two language models after post hoc exclusion of a third; agreement demonstrates reliability rather than clinical validity, and the harsher-tier rule may increase severe classifications without clinician calibration. The same model family contributed to automated error detection and severity grading, so those stages were not independent. Consultations were scripted and recorded under clean acoustic conditions, so robustness to noise was not assessed. The evaluated output was an unedited first draft from one proprietary system and configuration; no patient outcomes were measured. All authors are employees of the developer, which designed, funded and analysed the study, and the automated reviewer is an internal tool. Masking, multiple model families, sensitivity analyses and explicit reporting of adverse findings improve transparency but do not substitute for independent external replication.

## Conclusion

Across 20 paired simulated allied health consultations, AI-generated notes achieved modestly higher pooled PDQI-9 scores and lower total and omission error density than professional notes. The Critical+High contrast favoured AI directionally but was imprecise and compatible with no difference. The pooled quality estimate was sensitive to dietetics, and the small discipline-specific samples precluded confirmatory inference. The observed quality advantage was pronounced in dietetics, where source information was predominantly spoken, but small or absent in three examination-focused disciplines, where observations and measurements were incompletely represented in the audio. Transcript incompleteness, single primary quality ratings, post hoc rater-panel composition and automated severity grading further limit interpretation. Prospective, adequately powered, discipline-specific studies should evaluate clinician-reviewed notes using independent human adjudication and a multimodal reference standard encompassing spoken and observed clinical information.

## Supporting information

DECIDE-AI Checklist

## Acknowledgements

None

## Author contributions

Henry Bergman: Conceptualization, Methodology, Formal analysis, Investigation, Data curation, Writing - original draft, Supervision, Project administration. HB is the guarantor.

Vivian Liu: Methodology, Validation, Project administration, Writing - review & editing.

Sarah Ali: Methodology, Validation, Project administration, Writing - review & editing.

## Ethical considerations

All participants provided written informed consent. The Health Research Authority decision tool indicated that NHS Research Ethics Committee review was not required for the UK site; it does not constitute approval or a waiver by an institutional research ethics committee.^24^

## Consent to participate

All participants provided written informed consent to participate and for the research use of the resulting notes and recordings.

## Consent for publication

Not applicable. The manuscript contains no identifiable information relating to an individual participant.

## Data availability

Data supporting the reported findings are presented in the article and Supplementary Materials. The study-specific severity rubric, model configurations, false-positive-screen categories and decision rules are reported in the supplement. Verbatim prompts, item-level evaluation datasets, source scripts and analysis code are not publicly available because they contain proprietary or commercially sensitive material. Requests may be directed to the corresponding author and will be considered subject to participant consent, confidentiality, intellectual-property, data-governance and commercial requirements.

## References

1. Hassan H, Zipursky AR, Rabbani N, et al. Clinical implementation of artificial intelligence scribes in health care: a systematic review. Appl Clin Inform. 2025;16(4):1121–1135. doi:10.1055/a-2597-2017.

2. Lukac PJ, Turner W, Vangala S, et al. Ambient AI scribes in clinical practice: a randomized trial. NEJM AI. 2025. doi:10.1056/AIoa2501000.

3. Shah SJ, Devon-Sand A, Ma SP, et al. Ambient artificial intelligence scribes: physician burnout and perspectives on usability and documentation burden. J Am Med Inform Assoc. 2025;32(2):375–380. doi:10.1093/jamia/ocae295.

4. Topaz M, Peltonen LM, Zhang Z. Beyond human ears: navigating the uncharted risks of AI scribes in clinical practice. NPJ Digit Med. 2025;8(1):569. doi:10.1038/s41746-025-01895-6.

5. Coiera E, Fraile-Navarro D. AI scribes: are we measuring what matters? JMIR Med Inform. 2026;14:e89337. doi:10.2196/89337.

6. Ryan L, Hattingh L, Wall D, et al. Acceptance of AI scribes within hospital allied health settings: a mixed methods study. Digit Health. 2026;12:1–16. doi:10.1177/20552076261437234.

7. Ryan L, Hattingh L. Artificial intelligence medical scribes in allied health: a solution in search of evidence? Aust Health Rev. 2025;49:AH25064. doi:10.1071/AH25064.

8. Pain T, Kingston G, Askern J, et al. How are allied health notes used for inpatient care and clinical decision-making? A qualitative exploration of the views of doctors, nurses and allied health professionals. Health Inf Manag J. 2017;46(1):23–31. doi:10.1177/1833358316664451.

9. Schwartz-Dillard J, Ng T, Villegas J, et al. Electronic documentation burden among outpatient rehabilitation therapists: a qualitative descriptive study and quality improvement initiative. J Am Med Inform Assoc. 2024;31(10):2347–2355. doi:10.1093/jamia/ocae192.

10. Dahlberg A, Käenniemi T, Winther-Jensen T, et al. Measuring the quality of AI-generated clinical notes: a systematic review and experimental benchmark of evaluation methods. Artif Intell Med. 2026;177:103421. doi:10.1016/j.artmed.2026.103421.

11. Zhou L, Blackley SV, Kowalski L, et al. Analysis of errors in dictated clinical documents assisted by speech recognition software and professional transcriptionists. JAMA Netw Open. 2018;1(3):e180530. doi:10.1001/jamanetworkopen.2018.0530.

12. Asgari E, Montaña-Brown N, Dubois M, et al. A framework to assess clinical safety and hallucination rates of LLMs for medical text summarisation. NPJ Digit Med. 2025;8(1):274. doi:10.1038/s41746-025-01670-7.

13. Taylor SL, Jost M, MacDonald S, et al. Quality of clinical notes created by ambient listening generative AI: pragmatic prospective pilot study. JMIR Med Inform. 2026;14:e86474. doi:10.2196/86474.

14. Palm E, Manikantan A, Mahal H, et al. Assessing the quality of AI-generated clinical notes: validated evaluation of a large language model ambient scribe. Front Artif Intell. 2025;8:1691499. doi:10.3389/frai.2025.1691499.

15. Van Veen D, Van Uden C, Blankemeier L, et al. Adapted large language models can outperform medical experts in clinical text summarization. Nat Med. 2024;30(4):1134–1142. doi:10.1038/s41591-024-02855-5.

16. Vasey B, Nagendran M, Campbell B, et al. Reporting guideline for the early stage clinical evaluation of decision support systems driven by artificial intelligence: DECIDE-AI. BMJ. 2022;377:e070904. doi:10.1136/bmj-2022-070904.

17. Stetson PD, Bakken S, Wrenn JO, et al. Assessing electronic note quality using the Physician Documentation Quality Instrument (PDQI-9). Appl Clin Inform. 2012;3(2):164–174. doi:10.4338/ACI-2011-11-RA-0070.

18. International Organization for Standardization. ISO 14971:2019. Medical devices: application of risk management to medical devices. Geneva: ISO; 2019.

19. Feinstein AR, Cicchetti DV. High agreement but low kappa: I. The problems of two paradoxes. J Clin Epidemiol. 1990;43(6):543–549. doi:10.1016/0895-4356(90)90158-L.

20. Feinstein AR, Cicchetti DV. High agreement but low kappa: II. Resolving the paradoxes. J Clin Epidemiol. 1990;43(6):551–558. doi:10.1016/0895-4356(90)90159-M.

21. Landis JR, Koch GG. The measurement of observer agreement for categorical data. Biometrics. 1977;33(1):159–174. doi:10.2307/2529310.

22. Bergman H, Liu V, Austin B, et al. Quality, consistency, and clinical safety of AI-generated versus clinician-written clinical notes: a multi-country paired simulation study. medRxiv 2026.08.18.26360701. doi:10.64898/2026.08.18.26360701.

23. Norman GR, Sloan JA, Wyrwich KW. Interpretation of changes in health-related quality of life: the remarkable universality of half a standard deviation. Med Care. 2003;41(5):582–592. doi:10.1097/00005650-200305000-00004.

24. Health Research Authority. Do I need NHS REC review? HRA/MRC decision tool. https://www.hra-decisiontools.org.uk/ethics/ (accessed 12 August 2026).

25. Morey J, Jones D, Walker L, et al. Ambient artificial intelligence vs human scribes in the emergency department. Ann Emerg Med. 2026;87(5):561–568. doi:10.1016/j.annemergmed.2025.10.006.

26. Croxford E, Gao Y, First E, et al. Evaluating clinical AI summaries with large language models as judges. NPJ Digit Med. 2025;8(1):640. doi:10.1038/s41746-025-02005-2.

27. Genovese A, Hegstrom L, Prabha S, et al. Artificial authority: the promise and perils of LLM judges in healthcare. Bioengineering (Basel). 2026;13(1):108. doi:10.3390/bioengineering13010108.

28. Gładysz M, Fiumedinisi F, Burn F, et al. AI-assisted medical documentation in a multilingual Swiss health care system: proof-of-concept study. JMIR AI. 2026;5:e77351. doi:10.2196/77351.

29. Walker KJ, Wang A, Dunlop W, et al. The 9-item Physician Documentation Quality Instrument (PDQI-9) score is not useful in evaluating EMR (scribe) note quality in emergency medicine. Appl Clin Inform. 2017;8(3):981–993. doi:10.4338/ACI2017050080.

30. Mokkink LB, de Vet HCW, Prinsen CAC, et al. COSMIN Risk of Bias checklist for systematic reviews of Patient-Reported Outcome Measures. Qual Life Res. 2018;27(5):1171–1179. doi:10.1007/s11136-017-1765-4.

31. Sorin V, Klang E. AI scribe safety: measuring what happens after signing. JMIR Med Inform. 2026;14:e103162. doi:10.2196/103162.

32. Atiku S, Olakotan O. An evaluation of AI-generated clinical notes in the OpenNotes era: a thematic analysis of clinician discourse. J Eval Clin Pract. 2026;32(5):e70516. doi:10.1111/jep.70516.

