## Supplementary File for "Quality and transcript-grounded clinical-risk error burden of AI-generated vs allied health professional clinical notes: a paired simulation study across four disciplines"

### Contents


| Item | Title |
|---|---|
| Table S1 | PDQI-9 item profile |
| Table S2 | Sensitivity of pooled quality result |
| Table S3 | Error burden by discipline |
| Table S4 | Per-note severe-error counts |
| Table S5 | Three-model severity marginals |
| Table S6 | Severe-error register |
| Tables S7–S8 | Error type by discipline |
| Table S9 | Human unsupported-statement flags by discipline |
| Table S10 | Panel reliability and four-tier κ |
| Tables S11–S12 | False-positive screens |
| Table S13 | Targeted screen of Critical + High errors |
| Table S14 | Companion physician comparator (medRxiv preprint) |
| Table S15 | Error composition per note by author |
| Table S16 | Inventory of analysis outputs underlying reported results |
| Table S17 | Bayesian model specification and priors |
| Table S18 | Convergence diagnostics |
| Table S19 | Posterior predictive checks |
| Table S20 | Sensitivity of the Bayesian note-type estimate |
| Table S21 | Evaluator and note-writer nesting |
| Table S22 | Sample-size sensitivity and planning values |

| | |
|---|---|
| Table S23 | Precision of error-density contrasts |
| Table S24 | Evaluation pipeline and model configuration |
| Figure S1 | Unsupported-statement and omission density by discipline and note author |


#### Table S1. PDQI-9 item profile

PDQI-9 item profile, AI versus professional (n=20 consultations).

| Item | AI | Professional | Δ | dz | p (raw) | p (BH) |
|---|---|---|---|---|---|---|
| Comprehensible | 4.40 | 3.80 | +0.60 | 0.49 | 0.038 | 0.086 |
| Accurate | 4.35 | 3.80 | +0.55 | 0.52 | 0.032 | 0.086 |
| Internally consistent | 4.40 | 3.95 | +0.45 | 0.74 | 0.007 | 0.060 |
| Up-to-date | 4.35 | 3.95 | +0.40 | 0.53 | 0.033 | 0.086 |
| Organised | 4.25 | 3.85 | +0.40 | 0.31 | 0.206 | 0.309 |
| Useful | 4.20 | 3.90 | +0.30 | 0.32 | 0.157 | 0.283 |
| Synthesized | 4.10 | 3.80 | +0.30 | 0.23 | 0.376 | 0.376 |
| Thorough | 3.95 | 3.75 | +0.20 | 0.17 | 0.335 | 0.376 |
| Succinct | 4.45 | 4.25 | +0.20 | 0.24 | 0.285 | 0.366 |

#### Table S2. Sensitivity of the pooled quality result

| Analysis | n | Δ | 95% CI | Wilcoxon p |
|---|---|---|---|---|
| All four disciplines | 20 | +3.40 | +0.01 to +6.79 | 0.042 |
| Excluding dietetics | 15 | +0.87 | −2.35 to +4.08 | 0.489 |
| Excluding the two largest paired differences | 18 | +2.00 | −0.93 to +4.93 | 0.129 |

#### Table S3. Error burden by discipline

Automated reviewer; five notes per group per discipline.

| Discipline | Errors/note (AI) | Errors/note (Prof.) | C+H/note (AI) | C+H/note (Prof.) |
|---|---|---|---|---|

| | | | | |
|---|---|---|---|---|
| Dietetics | 1.60 | 6.20 | 0.40 | 0.20 |
| Physiotherapy | 2.60 | 9.00 | 0.40 | 1.20 |
| Occupational Therapy | 3.40 | 10.20 | 0.20 | 0.40 |
| Speech and Language Therapy | 5.80 | 8.80 | 0.40 | 1.20 |

**Table S4. Per-note Critical + High error counts**

Automated-reviewer distribution underlying the densities of 0.35 per AI note and 0.75 per professional note. Rows include notes with at least one severe error.

*Table S4a. AI notes*

AI (Heidi): 7 severe errors across 7 of 20 notes (density 0.35 per note).

| Consultation | Discipline | Severe errors |
|---|---|---|
| AH-SES-002 | Dietetics | 1 |
| AH-SES-004 | Dietetics | 1 |
| AH-SES-008 | Occupational Therapy | 1 |
| AH-SES-011 | Physiotherapy | 1 |
| AH-SES-012 | Physiotherapy | 1 |
| AH-SES-016 | Speech and Language Therapy | 1 |
| AH-SES-017 | Speech and Language Therapy | 1 |

*Table S4b. Professional notes*

Professional: 15 severe errors across 6 of 20 notes (density 0.75 per note).

| Consultation | Discipline | Severe errors |
|---|---|---|
| AH-SES-011 | Physiotherapy | 5 |
| AH-SES-017 | Speech and Language Therapy | 4 |
| AH-SES-006 | Occupational Therapy | 2 |
| AH-SES-016 | Speech and Language Therapy | 2 |
| AH-SES-005 | Dietetics | 1 |

| | | |
|---|---|---|
| AH-SES-014 | Physiotherapy | 1 |

**Table S5. Per-model severity marginals from the initial three-model panel**

Grades assigned to the same 238 errors before the consensus rule. The exclusion of Gemini 2.5 Pro followed inspection of these outputs and is therefore post hoc.

| Model | Low | Moderate | High | Critical | % Critical+High | Matches consensus |
|---|---|---|---|---|---|---|
| gpt-5.5 | 92 | 126 | 17 | 3 | 8.4% | 80.3% |
| claude-opus-4-8 | 50 | 178 | 9 | 1 | 4.2% | 84.5% |
| gemini-2.5-pro | 32 | 65 | 104 | 37 | 59.2% | 34.9% |

**Table S6. Severe-error register**

The analysis identified 22 Critical + High errors in the automated-reviewer arm and six in the professional-adjudication arm. The register below retains the reported grades and cited evidence. These are simulated-risk classifications, not observed patient harms.

Calibrated automated reviewer: 22 severe errors

AH-SES-016: Speech and Language Therapy: Professional note: CRITICAL (S×L 16): unsupported statement; models disagreed, harsher grade taken

Admission diagnosis documented as LMCA despite the transcript only supporting a general stroke context, not a left MCA diagnosis.

{"note_claim":"Admitted with LMCA","transcript_evidence":"Transcript refers to stroke context and aphasia/arm weakness, but does not specify left MCA or LMCA admission diagnosis."}

AH-SES-002: Dietetics: AI (Heidi) note: HIGH (S×L 12): unsupported statement; models disagreed, harsher grade taken

The note states the patient's father had bilateral toe amputations, but the transcript only says he 'lost two toes' and does not specify bilateral involvement.

{"note_claim":"Father had type 2 diabetes with poor management, resulting in bilateral toe amputations.","transcript_excerpt":"My dad had it. He didn't manage it well. Lost two toes."}

AH-SES-004: Dietetics: AI (Heidi) note: HIGH (S×L 12): unsupported statement; models disagreed, harsher grade taken

The note lists 'spicy fruit' as a known trigger, but the transcript states 'very spicy food'.

{"note_claim":"Known food triggers: corn (on the cob, kernels), spicy fruit, alcohol, raw onion.","transcript_excerpt":"Corn. Like— corn on the cob or kernels. Always. And very spicy food. ...

Alcohol ... And um— raw onion sometimes.","issue":"The note changes 'very spicy food' to 'spicy fruit'."}

AH-SES-005: Dietetics: Professional note: HIGH (S×L 12): unsupported statement; both models agreed

The note states nutritional intervention is being undertaken prior to a phosphate binder, whereas the transcript discusses considering a potassium binder if dietary measures do not lower potassium.

{"note_claim":"nutritional intervention prior to phosphate binder","transcript_excerpt":"Then we'd look at a medication called a potassium binder that helps remove it in the gut. But let's try the diet first."}

AH-SES-006: Occupational Therapy: Professional note: HIGH (S×L 12): unsupported statement; models disagreed, harsher grade taken

The note attributes the patient's right-sided weakness and poor motor control to a neurological condition, but no neurological diagnosis or condition is stated in the transcript.

{"note_claim":"secondary to neurological condition","transcript_excerpt":"PATIENT: This hand is— the right hand is not- ... Getting in and out of a bath with a weak right arm and leg needs some adaptations.","issue":"Right-sided weakness is described, but no neurological condition or diagnosis is named."}

AH-SES-006: Occupational Therapy: Professional note: HIGH (S×L 12): unsupported statement; models disagreed, harsher grade taken

The nursing plan instructs staff to provide assistance of one for all ADLs, but the transcript does not establish that level of assistance for all ADLs.

{"note_claim":"provide AoI for all ADLs","transcript_excerpt":"Patient required verbal cueing during washing task; patient reported nurses help with dressing and that buttons are difficult. No statement indicates AoI is required for all ADLs.","issue":"Assistance of one for all ADLs is not supported by the transcript."}

AH-SES-008: Occupational Therapy: AI (Heidi) note: HIGH (S×L 12): omission; models disagreed, harsher grade taken

Current safety measure of locking the kitchen due to gas cooker risk is omitted.

{"transcript_excerpt":"He left it on twice. We've been— I've been locking the kitchen.","note_context":"Note documents gas cooker safety concerns and that wife does most cooking, but does not document kitchen locking."}

AH-SES-011: Physiotherapy: AI (Heidi) note: HIGH (S×L 12): unsupported statement; models disagreed, harsher grade taken

The note states the patient had initial sharp pain during sit-to-stand, but the patient explicitly denied sharp pain.

{"note_claim":"Initial sharp pain","transcript_excerpt":"HEALTHCARE PROFESSIONAL: Is any of it sharp? Like really sharp? PATIENT: No, more— dull. Achy. Just a lot of it."}

AH-SES-011: Physiotherapy: Professional note: HIGH (S×L 12): unsupported statement; models disagreed, harsher grade taken

Daughter documented as next of kin without source support.

{"note_claim":"Daughter as NOK","transcript_evidence":"Patient says her daughter stayed over and will stay with her for the first week, but no next-of-kin designation is discussed."}

AH-SES-011: Physiotherapy: Professional note: HIGH (S×L 12): unsupported statement; models disagreed, harsher grade taken

Sitting on edge of bed documented as independent despite transcript indicating physiotherapist assistance was planned or provided.

{"note_claim":"SOEOB independent","transcript_evidence":"Physiotherapist: 'I'll help you swing your legs round'; patient then sits up at the edge of the bed."}

AH-SES-011: Physiotherapy: Professional note: HIGH (S×L 12): unsupported statement; models disagreed, harsher grade taken

Sit-to-stand documented as independent without clear transcript support.

{"note_claim":"STS Ind","transcript_evidence":"Physiotherapist gave instructions and was present with the frame; the transcript does not state the patient stood independently."}

AH-SES-011: Physiotherapy: Professional note: HIGH (S×L 12): unsupported statement; models disagreed, harsher grade taken

Second documentation of sit-to-stand as independent is unsupported by the transcript.

{"note_claim":"STS-Ind","transcript_evidence":"The transcript documents the patient standing with coaching and a frame, but does not explicitly document independent sit-to-stand."}

AH-SES-011: Physiotherapy: Professional note: HIGH (S×L 12): unsupported statement; models disagreed, harsher grade taken

Sitting into the chair documented as independent without clear transcript support.

{"note_claim":"Sat in chair independently","transcript_evidence":"Physiotherapist instructed: 'Now lower yourself down slowly. Hands on the armrests, don't drop.' The transcript does not state independent performance."}

AH-SES-012: Physiotherapy: AI (Heidi) note: HIGH (S×L 12): unsupported statement; both models agreed

The note documents muffled sensation bilaterally, but the transcript indicates the patient endorsed both heaviness and muffled sensation in the right leg, not bilateral symptoms.

{"claim":"Right leg feels heavy, as if underwater; muffled sensation bilaterally","transcript_evidence":["PATIENT: My leg feels like it's— sort of, underwater.","HEALTHCARE PROFESSIONAL: Heavy? Muffled sensation?","PATIENT: Both. Kind of, yeah."],"issue":"'Both' refers to heaviness and muffled sensation, not both sides of the body."}

AH-SES-014: Physiotherapy: Professional note: HIGH (S×L 12): omission; models disagreed, harsher grade taken

The note omits key activity restrictions: the cuff should not be significantly loaded, with no lifting and no carrying bags.

{"note_text":"advice given activity - no overhead work currently","transcript_excerpt":"At six weeks we start introducing more active movement, but we're still not loading the cuff significantly. No lifting, no carrying bags."}

AH-SES-016: Speech and Language Therapy: AI (Heidi) note: HIGH (S×L 12): unsupported statement; models disagreed, harsher grade taken

The note specifies a left MCA stroke, but the transcript only establishes that the patient is on a stroke unit and has had a stroke-related presentation.

{"note_claim":"Left MCA stroke","transcript_excerpts":["I'm one of the speech and language therapists on the stroke unit.","after this kind of stroke","my arm— gone","The right one isn't— he can't grip yet"],"issue":"Transcript supports stroke with aphasia/right-sided impairment but does not specify left MCA territory."}

AH-SES-016: Speech and Language Therapy: Professional note: HIGH (S×L 12): omission; models disagreed, harsher grade taken

Reading difficulty was incompletely documented: the patient reported he could read only some of a letter and that reading was very slow.

{"missing_detail":"slow. Very slow","transcript_excerpt":"HCP: 'And how was that— could you read it?' Patient: 'Some. Some— but— slow. Very slow.'","note_context":"The note states receptive language is a strength and that the patient is able to understand verbal and written language, but does not capture the reduced speed/partial reading ability."}

AH-SES-017: Speech and Language Therapy: AI (Heidi) note: HIGH (S×L 12): omission; both models agreed

The note does not document key PEG timing counselling that PEG becomes higher risk, and may become impossible, once FVC drops below a threshold.

{"transcript_excerpt":"The procedure is much safer when respiratory function is still good... Once it drops below a certain threshold, the procedure becomes higher-risk and sometimes can't be done at all.","note_excerpt":"FVC: 64% (respiratory review 1 month ago): within acceptable window for PEG procedure."}

AH-SES-017: Speech and Language Therapy: Professional note: HIGH (S×L 12): omission; models disagreed, harsher grade taken

The note omits the explicit counselling that the chest infection was very likely due to aspiration.

{"transcript_excerpt":"There's been a chest infection, which is very likely to be from food or fluid going the wrong way— what we call aspiration.","note_content":"The note records chest infection requiring antibiotics but does not explicitly document suspected aspiration."}

AH-SES-017: Speech and Language Therapy: Professional note: HIGH (S×L 12): omission; both models agreed

The note omits the recent respiratory function information, including the reported FVC of approximately 64%, which was used in counselling about PEG timing.

{"transcript_excerpt":"HUSBAND: A month ago. They said the FVC was— sixty something? Sixty-four? HEALTHCARE PROFESSIONAL: That's still in a good window for the procedure. But it's a window that closes...","note_content":"No FVC value or respiratory timing context is documented."}

AH-SES-017: Speech and Language Therapy: Professional note: HIGH (S×L 12): omission; models disagreed, harsher grade taken

The note omits significant mealtime fatigue and the related advice to adjust meal timing or use smaller, more frequent meals.

{"transcript_excerpt":"HEALTHCARE PROFESSIONAL: Are you noticing fatigue during meals? PATIENT: Massively. By the end of dinner I'm— I'm exhausted. HEALTHCARE PROFESSIONAL: Fatigue makes the swallow less safe... bigger meal at lunch... lighter in the evening. Or smaller, more frequent meals.","note_content":"No mealtime fatigue or meal-timing advice is documented."}

AH-SES-017: Speech and Language Therapy: Professional note: HIGH (S×L 12): omission; models disagreed, harsher grade taken

The note omits specific swallow safety strategies advised, including small spoons, upright positioning, no talking while eating, chin-tuck swallow, and small amounts at a time.

{"transcript_excerpt":"There are a few things that help— small spoons, sitting fully upright, no talking while eating, a chin-tuck swallow which I'll show you. And small amounts at a time.","note_content":"The plan records IDDSI Level 4 and Level 3 but not these swallow safety strategies."}

Professional adjudication: 6 severe errors

AH-SES-005: Dietetics: Professional note: CRITICAL (S×L 16): unsupported statement; models disagreed, harsher grade taken

The scenario involves a potassium binder rather than a phosphate binder.

Transcript: “Then we'd look at a medication called a potassium binder that helps remove it in the gut.”

AH-SES-008: Occupational Therapy: Professional note: HIGH (S×L 12): omission; models disagreed, harsher grade taken

Important evaluation information is missing from the objective assessment.

Transcript: “WIFE: Yes. He left it on twice. We've been— I've been locking the kitchen.”

AH-SES-017: Speech and Language Therapy: Professional note: HIGH (S×L 12): omission; models disagreed, harsher grade taken

The ongoing plan is missing.

Transcript: “HEALTHCARE PROFESSIONAL: I'll leave you a written sheet with all of this. And I'll book the joint appointment. I think you should take the next two weeks to read about the PEG, talk to each other, talk to the kids if you want to. ... I'll call tomorrow with the joint appointment time. If anything happens before then— more choking, more chest symptoms, anything that frightens you— please ring the team line and someone will speak to you.”

AH-SES-017: Speech and Language Therapy: Professional note: HIGH (S×L 12): omission; models disagreed, harsher grade taken

Additional clinical steps are missing.

Transcript: “HEALTHCARE PROFESSIONAL: Most people do at first. There are a few things that help— small spoons, sitting fully upright, no talking while eating, a chin-tuck swallow which I'll show you. And small amounts at a time. Are you noticing fatigue during meals? ... One other thing— how often do you do mouth care? ... Can we go up to four times? Including before bed and after every meal? Bacteria in the mouth that get aspirated cause more chest infections than the food itself, often. Good mouth care is one of the strongest protections.”

AH-SES-017: Speech and Language Therapy: Professional note: HIGH (S×L 12): omission; models disagreed, harsher grade taken

Additional patient steps are missing.

Transcript: “There are a few things that help— small spoons, sitting fully upright, no talking while eating, a chin-tuck swallow which I'll show you. And small amounts at a time.”

AH-SES-017: Speech and Language Therapy: Professional note: HIGH (S×L 12): omission; models disagreed, harsher grade taken

Salient information is not fully included in the body of the description.

Transcript: “HEALTHCARE PROFESSIONAL: This is the part where timing matters. The procedure is much safer when respiratory function is still good. There's a measure— forced vital capacity— that the team will monitor. Once it drops below a certain threshold, the procedure becomes higher-risk and sometimes can't be done at all. ... HUSBAND: A month ago. They said the FVC was— sixty something? Sixty-four?”

**Table S7. Error type by discipline, automated reviewer**

Errors per note; five notes per group per discipline.

| Discipline | AI unsupported | AI omission | Prof. unsupported | Prof. omission |
|---|---|---|---|---|
| Dietetics | 0.80 | 0.80 | 1.40 | 4.80 |
| Physiotherapy | 1.40 | 1.20 | 3.80 | 5.20 |
| Occupational Therapy | 0.60 | 2.80 | 4.60 | 5.60 |
| Speech and Language Therapy | 4.80 | 1.00 | 3.60 | 5.20 |

**Table S8. Error type by discipline, professional adjudication**

Errors per note; five notes per group per discipline.

| Discipline | AI unsupported | AI omission | Prof. unsupported | Prof. omission |
|---|---|---|---|---|
| Dietetics | 0.80 | 1.20 | 0.80 | 3.00 |
| Physiotherapy | 0.20 | 0.00 | 0.20 | 0.00 |
| Occupational Therapy | 0.00 | 1.20 | 0.20 | 1.00 |
| Speech and Language Therapy | 0.00 | 0.40 | 0.20 | 0.80 |

**Table S9. Human unsupported-statement flags by discipline**

The tables below present the available unsupported-statement flags by discipline. No independent coding of examination-related categories was undertaken, so the examples should not be interpreted as category frequencies.

*Table S9a. Dietetics*

| Consultation | Tier | Flag | Evidence cited |
|---|---|---|---|
| AH-SES-001 | MEDIUM | The note states the patient was 'Admitted for NOK', which is not supported by the transcript and does not reflect the stated admission context of a fall/hip operation. | {"note_claim":"Admitted for NOK","transcript_evidence":"The transcript refers to 'before the fall' and 'After a hip operation' but does not state admission for NOK."} |
| AH-SES-001 | LOW | The note records banana as the patient's favourite supplement flavour, but the transcript only indicates the patient's husband considered banana the 'least bad'. | {"note_claim":"banana her favourite","transcript_evidence":"Patient: 'Banana. He always said banana was the least bad.' Dietitian: 'I'll start with banana.'"} |

| | | | |
|---|---|---|---|
| AH-SES-002 | MEDIUM | Alcohol intake is documented with an unsupported and ambiguous frequency/unit: '3-4 beers over the weekend/per day' rather than the stated weekend pints. | {"note_claim":"Alcohol: 3-4 beers over the weekend/per day","transcript_excerpt":"Beer at weekends— maybe three or four pints Friday, Saturday."} |
| AH-SES-002 | LOW | The note states 'she needs to work around' workplace communal biscuits, but the transcript indicates this is the patient's issue, not his wife's. | {"note_claim":"Issues with large amounts of processed food like biscuits in working communal areas that she needs to work around","transcript_excerpt":"The biscuits at work are goi[ng...]"} |
| AH-SES-004 | MEDIUM | The note inaccurately states the patient had 4 days of no eating prior to admission, conflating the 4 days on IV fluids in hospital with the pre-admission period of approximately 1 week of minimal intake. | {"note_claim":"4 days no eating prior to admission","transcript_evidence":["you've been on the drip for— it's four days now, isn't it?","For about a week I was basically just— wat[ching...]"]} |
| AH-SES-005 | MEDIUM | The note labels the referral as a pre-dialysis consultation, but dialysis or pre-dialysis status is not stated in the transcript. | {"note_claim":"Ref: Pre dialysis consultation","transcript_evidence":"transcript discusses advancing kidney disease, raised potassium and phosphate, and renal dietetic input, b[ut...]"} |
| AH-SES-005 | HIGH | The note states nutritional intervention is being undertaken prior to a phosphate binder, whereas the transcript discusses considering a potassium binder if dietary measures do not lower potassium. | {"note_claim":"nutritional intervention prior to phosphate binder","transcript_excerpt":"Then we'd look at a medication called a potassium binder that helps remove it in the gut. B[ut let's try the diet first.]"} |

*Table S9b. Physiotherapy*

| Consultation | Tier | Flag | Evidence cited |
|---|---|---|---|
| AH-SES-011 | MEDIUM | Encounter date documented without support in the transcript. | {"note_claim":"@8.05.2026","transcript_evidence":"The transcript does not state an encounter date."} |
| AH-SES-011 | MEDIUM | Encounter time documented without support in the transcript. | {"note_claim":"Seen at 11:30am","transcript_evidence":"The transcript starts with 'Morning' but gives no exact time."} |
| AH-SES-011 | HIGH | Daughter documented as next of kin without source support. | {"note_claim":"Daughter as NOK","transcript_evidence":"Patient says her daughter stayed over and will stay with her for the first week, but no next-of-kin designation is discussed."} |
| AH-SES-011 | HIGH | Sitting on edge of bed documented as independent despite transcript indicating physiotherapist assistance was planned/provided. | {"note_claim":"SOEOB independent","transcript_evidence":"Physiotherapist: 'I'll help you swing your legs round'; patient then sits up at the edge of the bed."} |

| | | | |
|---|---|---|---|
| AH-SES-011 | HIGH | Sit-to-stand documented as independent without clear transcript support. | {"note_claim":"STS Ind","transcript_evidence":"Physiotherapist gave instructions and was present with the frame; the transcript does not state the patient stood independently."} |
| AH-SES-011 | MEDIUM | Dizziness is documented in association with standing, but it occurred after sitting on the edge of the bed and resolved before standing. | {"note_claim":"Standing//dizzy","transcript_evidence":"After sitting on edge of bed, patient says 'Dizzy. Just for a second'; after resting she says 'Better now' before standing."} |
| AH-SES-011 | HIGH | Second documentation of sit-to-stand as independent is unsupported by the transcript. | {"note_claim":"STS-Ind","transcript_evidence":"The transcript documents the patient standing with coaching and a frame, but does not explicitly document independent sit-to-stand."} |
| AH-SES-011 | HIGH | Sitting into the chair documented as independent without clear transcript support. | {"note_claim":"Sat in chair independently","transcript_evidence":"Physiotherapist instructed: 'Now lower yourself down slowly. Hands on the armrests, don't drop.' The transcript does not state independent performance."} |

*Table S9c. Occupational therapy*

| Consultation | Tier | Flag | Evidence cited |
|---|---|---|---|
| AH-SES-006 | MEDIUM | The note states the patient can independently don/doff Velcro shoes, but the transcript only supports that she usually wears slip-on/Velcro shoes and had done so before admission. | {"note_claim":"patient reported that patient can independently don/doff Velcro shoes","transcript_excerpt":"PATIENT: I usually wear— I have those slip-on ones. Velcro. My feet..."} |
| AH-SES-006 | MEDIUM | The note documents the patient's goal as showering independently, but the patient actually stated she wanted a proper wash independently and reported having a bath with no shower at home. | {"note_claim":"Patient reported that she wants to be able to shower on her own","transcript_excerpt":"PATIENT: Have a proper wash. By myself. HEALTHCARE PROFESSIONAL: And the b[ath...]"} |
| AH-SES-006 | MEDIUM | The note states the patient is 67 years old, but no age is provided in the transcript. | {"note_claim":"Patient is 67 yo female","transcript_excerpt":"No patient age is stated in the transcript.","issue":"Age is unsupported by the source transcript."} |
| AH-SES-006 | MEDIUM | The analysis states the patient requires assistance of one to complete ADLs, but the transcript documents cueing/support during assessment and some nursing help, not a formal AoI requirement. | {"note_claim":"requiring AoI to complete ADLs","transcript_excerpt":"HEALTHCARE PROFESSIONAL: Can you try using your left hand to guide the right? ... Do you want to try one-handed..."} |

| | | | |
|---|---|---|---|
| AH-SES-006 | HIGH | The note attributes the patient's right-sided weakness and poor motor control to a neurological condition, but no neurological diagnosis or condition is stated in the transcript. | {"note_claim":"secondary to neurological condition","transcript_excerpt":"PATIENT: This hand is— the right hand is not- ... Getting in and out of a bath with a weak right arm and l[eg...]"} |
| AH-SES-006 | HIGH | The nursing plan instructs staff to provide assistance of one for all ADLs, but the transcript does not establish that level of assistance for all ADLs. | {"note_claim":"provide AoI for all ADLs","transcript_excerpt":"Patient required verbal cueing during washing task. Patient reported nurses help with dressing and that buttons are d[ifficult...]"} |
| AH-SES-007 | MEDIUM | The note overstates the duration of being in bed by documenting that the patient spent much of both Wednesday and Thursday in bed, whereas the transcript supports most of Wednesday and only a patchy Thursday. | {"note_claim":"She reported that she had to spend much of Wednesday and Thursday in bed","transcript_excerpt":"PATIENT: Most of Wednesday. Thursday was patchy. Friday I started to..."} |
| AH-SES-007 | MEDIUM | The note states the patient completed a large number of tasks, but the transcript only states that she overfilled her activity schedule and tried to do too much. | {"note_claim":"Gemma had independently planned her week and completed a large number of tasks","transcript_excerpt":"PATIENT: I tried to do it myself and I- I filled it in with to[o much...]"} |

*Table S9d. Speech and language therapy*

| Consultation | Tier | Flag | Evidence cited |
|---|---|---|---|
| AH-SES-016 | MEDIUM | Patient age documented as 67 years old without support in the transcript. | {"note_claim":"67y/o","transcript_evidence":"The transcript does not state the patient's age."} |
| AH-SES-016 | CRITICAL | Admission diagnosis documented as LMCA despite the transcript only supporting a general stroke context, not a left MCA diagnosis. | {"note_claim":"Admitted with LMCA","transcript_evidence":"Transcript refers to stroke context and aphasia/arm weakness, but does not specify left MCA or LMCA admission diagnosis."} |
| AH-SES-016 | MEDIUM | Naming task score documented as 3/5 correct, but the transcript only shows three naming items attempted and all were ultimately named correctly. | {"note_claim":"Naming task: 3/5 correct","transcript_evidence":"Items documented: cup named; clock retrieved after delay/cue; window named. No five-item task is evidenced."} |
| AH-SES-016 | MEDIUM | Semantic errors documented during naming, but the transcript shows word-finding difficulty and circumlocution rather than a clear semantic naming error. | {"note_claim":"noted semantic errors","transcript_evidence":"Patient circumlocutes, e.g. 'time thing' before retrieving 'clock', and uses metaphorical [description]; no clear semantic..."} |
| AH-SES-016 | MEDIUM | Phonological cueing documented as beneficial, but no phonological cueing is recorded in the transcript. | {"note_claim":"Patient benefitted from phonological cueing","transcript_evidence":"The SLT provides time and a semantic cue ('It tells the time') but no initial sound, syllable..."} |

| | | | |
|---|---|---|---|
| AH-SES-016 | MEDIUM | The note states the patient needs daily therapy input, but the transcript only supports a planned review the next morning and brief practice exercises. | {"note_claim":"needs daily therapy input","transcript_evidence":"SLT says, 'I'll see you tomorrow morning' and 'Even five minutes here and there makes a difference'; no explicit ne[ed for daily input]"} |
| AH-SES-017 | MEDIUM | The note states that consent was obtained, but explicit consent is not present in the transcript. | {"note_claim":"Consent obtained","transcript_evidence":"The transcript begins with greetings and discussion of symptoms; no explicit consent request or confirmation is documented."} |
| AH-SES-017 | MEDIUM | The note attributes 'more frequent choking episodes' to the husband, but the transcript documents worsening symptoms and one severe episode rather than an explicit increase in frequency reported by him. | {"note_claim":"Husband advised of more frequent choking episodes","transcript_excerpt":"HUSBAND: She had a bad one on Sunday. With water. We thought- I thought I was going to have..."} |

#### Table S10. Severity-panel reliability

| Measure | Calibrated arm (n=238) | Professional arm (n=50) |
|---|---|---|
| Exact four-tier agreement | 66.0% | 58.0% |
| Agreement, severe versus not | 92.9% | 90.0% |
| Cohen's κ, severe versus not | 0.339 | 0.260 |
| Fleiss' κ, four-tier | 0.270 | 0.255 |

#### Table S11. False-positive screen for unsupported statements

| Arm | Flags | Genuine | Reasonable inference | Benign detail | False-positive rate |
|---|---|---|---|---|---|
| AI (Heidi) | 38 | 78.9% | 5 | 3 | 7.9% |
| Professional | 67 | 77.6% | 14 | 1 | 1.5% |

Counts in the middle columns are numbers of flags; percentages are shown only where the source table uses a percentage sign.

#### Table S12. False-positive screen for omissions

| Arm | Flags | Genuine | Appropriate concision | Not clinically significant | Present but reworded | False-positive rate |
|---|---|---|---|---|---|---|
| AI (Heidi) | 29 | 65.5% | 7 | 2 | 1 | 34.5% |

| | | | | | | |
|---|---|---|---|---|---|---|
| Professional | 104 | 80.8% | 1 | 7 | 2 | 19.2% |

Counts in the middle columns are numbers of flags; percentages are shown only where the source table uses a percentage sign.

#### Table S13. Targeted screen of the 22 Critical + High automated-reviewer errors

| Verdict | n |
|---|---|
| genuine | 16 |
| uncertain | 3 |
| reasonable inference | 2 |
| appropriate concision | 1 |

Targeted screening classified 16 flags as confirmed genuine errors: six in AI notes and 10 in professional notes, corresponding to 0.30 and 0.50 per note. These post-screen values are distinct from the pre-screen automated-reviewer densities of 0.35 and 0.75 per note.

#### Table S14. Allied health findings and companion physician-study preprint

| Measure | Allied health (n=20) | Physicians (n=385) |
|---|---|---|
| PDQI-9, AI | 38.45 | 40.63 |
| PDQI-9, human | 35.05 | 35.55 |
| Difference | +3.40 (dz 0.47) | +5.08 (dz 0.55) |
| Mixed-model AI advantage | +3.47 (94% ETI +0.66 to +6.30) | +5.11 (HDI +4.37 to +5.88) |
| Dispersion (SD, AI vs human) | 7.21 vs 7.19 (p=0.46) | 4.97 vs 7.76 ($p=4\times10^{-13}$) |
| “Poor” notes, AI vs human | 15% vs 40% | 5.7% vs 27.8% |
| Severe density, AI | 0.35 / note | 0.335 / note |
| Severe density, human | 0.75 / note | 1.271 / note |
| Severe-density ratio | 2.1x | 3.8x |
| Human detection sensitivity | 8.4% / 9.5% | ~12% |

#### Table S15. Error composition per note by author

Per-note joint error composition under the calibrated automated reviewer, with 20 notes per group. Values are the percentage of notes in each arm falling into each mutually exclusive category, as represented in the corresponding error-composition output. Rows sum to 100%.

| Author | No error found | Unsupported only | Omission only | Both |
|---|---|---|---|---|
| AI (Heidi) | 15% | 15% | 25% | 45% |
| Professional | 0% | 0% | 5% | 95% |

The categories are mutually exclusive and assigned per note. This table therefore describes how errors were distributed across notes and complements the per-note densities in Table 3 of the main manuscript, which describe the number of errors.

#### Table S16. Inventory of analysis outputs underlying reported results

Mapping between reported results and the analysis outputs that produced them. The pipeline was version-controlled and applied to a frozen data snapshot; each stage wrote a separate output, and severity scoring required a grade from every declared panel member.

| Analysis output | Contents |
|---|---|
| summary.json; step0*.csv | PDQI-9 quality chain (steps 03, 03b, 04, 05, 06, 07, 08, 10) |
| risk_allied_health.json | Calibrated-arm severity aggregate and reliability |
| risk_scores_allied_health.json | Per-error granular severity (238 errors, both raters) |
| risk_human_allied_health.json; risk_scores_human_allied_health.json | Professional-adjudication arm (50 errors) |
| calibration_allied_health.json; calibration_omission_allied_health.json | False-positive screens by error type |
| calibration_crithigh_allied_health.json | Targeted screen of the 22 Critical + High errors |
| confusion_allied_health.json | Detection sensitivity and precision, overall and by author |
| _ahp_3panel_evidence/ | Three-model panel output retained as evidence for the panel-composition decision |
| ahp_new_analyses results | Bayesian diagnostics, posterior predictive checks, sensitivity models, nesting, sample-size planning and paired-bootstrap intervals |
| diagnostic posterior draws | Convergence, effective sample sizes, divergent transitions and posterior predictive simulations |

#### *Additional analytical methods and sensitivity analyses*

All additional analyses used the same frozen dataset and cleaning path as the main analysis. The primary Bayesian model reproduced the reported note-type estimate of +3.47 points. The analyses below correct the interval label, document model diagnostics and assumptions, quantify precision for the safety co-primary outcome and describe the evaluation pipeline without disclosing proprietary prompts.

*Bayesian model specification*

The model was PDQI-9 total ~ note type + (1|consultation) + (1|evaluator) + (1|note writer), fitted to 40 notes from 20 consultations using a Gaussian likelihood and identity link. Each AI note carried the identifier of the professional who wrote the paired note, so its note-writer term functions as a clustering factor for the paired professional rather than a direct measure of AI writing skill. Software was bambi 0.18.0, PyMC 6.0.1 and ArviZ 1.1.0, with seed 20260518. Priors were bambi's default weakly informative priors, scaled to the observed outcome SD and not tuned to the study. The primary fit used two chains, 1,000 post-warm-up draws per chain, 1,000 tuning steps and target acceptance 0.90. A diagnostic refit used four chains, 2,000 post-warm-up draws per chain, 2,000 tuning steps and target acceptance 0.95. One empty incomplete record was excluded before analysis; all 40 analysed notes had complete PDQI-9 items and identifiers, giving 20 complete pairs with no imputation. The 94% interval was generated from the 3rd and 97th posterior percentiles; it was retained for reproducibility and was not used as a confirmatory decision threshold.

**Table S17. Bayesian model priors**

| Parameter | Prior |
|---|---|
| Intercept | Normal(36.75, 25.52) |
| Note type (professional versus AI) | Normal(0, 36.09) |
| Consultation, evaluator and note-writer intercepts | Normal(0, sigma-k); sigma-k ~ HalfNormal(25.52) |
| Residual SD | HalfStudentT(nu=4, 7.22) |

The priors were diffuse relative to the 9-45 PDQI-9 scale. The prior median for each random-effect SD was 17.3, compared with posterior medians of 1.1-5.4, indicating that the posterior estimates were principally informed by the observed data.

**Table S18. Convergence diagnostics**

| Fit | AI advantage | 94% ETI | 94% HDI | P(AI > professional) | Maximum R-hat | Minimum bulk/tail ESS | Divergences | E-BFMI |
|---|---|---|---|---|---|---|---|---|
| Primary: 2 x 1,000 | +3.47 | +0.66 to +6.30 | +0.61 to +6.17 | 0.989 | 1.01 | 334 / 547 | 4/2,000 (0.20%) | 0.77-0.82 |
| Diagnostic: 4 x 2,000 | +3.44 | +0.65 to +6.18 | +0.61 to +6.13 | 0.988 | 1.00 | 2,006 / 2,537 | 10/8,000 (0.13%) | 0.81-0.87 |

R-hat was at most 1.01 and all bulk effective sample sizes exceeded 300. The analysis code calculated the 3rd and 97th posterior percentiles, so +0.66 to +6.30 is a 94% equal-tailed interval rather than an HDI. The corresponding 94% HDI was +0.61 to +6.17. The small number of divergences did not change the note-type interpretation; zero-divergence sensitivity models gave similar estimates (Table S20).

**Table S19. Posterior predictive checks**

| Statistic | Observed | Replicated median (95% interval) | Posterior predictive p |
|---|---|---|---|
| Mean, AI | 38.45 | 38.42 (35.57 to 41.29) | 0.49 |
| Mean, professional | 35.05 | 35.07 (32.20 to 37.98) | 0.51 |
| SD, AI | 7.21 | 7.31 (5.19 to 9.77) | 0.54 |
| SD, professional | 7.19 | 7.43 (5.31 to 9.94) | 0.58 |
| Mean paired difference | 3.40 | 3.39 (-0.84 to 7.43) | 0.50 |
| SD of paired differences | 7.24 | 6.49 (4.19 to 9.77) | 0.31 |
| Proportion below 32 | 0.28 | 0.25 (0.15 to 0.38) | 0.49 |
| Minimum | 21 | 19.8 (11.5 to 25.8) | 0.37 |
| Maximum | 45 | 51.3 (46.3 to 59.1) | 0.995 |

The model reproduced the observed location, spread, paired difference and low-score tail. The main misfit was the bounded scale ceiling: six of 40 notes scored 45, whereas the Gaussian likelihood was unbounded and 13.7% of replicated scores exceeded 45. Residual normality was acceptable (Shapiro-Wilk p=0.19; skew -0.29), as was normality of paired differences (p=0.35). Residual variance was similar by arm (SD 4.07 versus 3.75; Levene p=0.71) and discipline (Levene p=0.15). Clustering by consultation, evaluator and writer was modelled explicitly.

### Table S20. Sensitivity of the Bayesian note-type estimate

| Specification | AI advantage | 94% ETI or 95% CI | P(AI > professional) | Divergences |
|---|---|---|---|---|
| Primary | +3.47 | +0.66 to +6.30 | 0.989 | 4 |
| Wide priors | +3.45 | +0.69 to +6.28 | 0.989 | 1 |
| Regularising priors | +3.19 | +0.49 to +5.86 | 0.986 | 0 |
| Student-t likelihood | +3.43 | +0.54 to +6.32 | 0.987 | 0 |
| Right-censored at 45 | +3.50 | +0.11 to +6.95 | 0.973 | 4 |
| Discipline fixed effect | +3.46 | +0.56 to +6.32 | 0.987 | 19 |
| Consultation random effect only | +3.39 | +0.08 to +6.80 | 0.973 | 0 |
| Frequentist REML, crossed components | +3.46 | 95% CI +0.83 to +6.09 | p=0.010 | Not applicable |

| | | | | |
|---|---|---|---|---|
| Frequentist REML, consultation only | +3.40 | 95% CI +0.23 to +6.57 | p=0.036 | Not applicable |

Across prior scales, likelihoods, ceiling handling and estimation frameworks, the point estimate remained between +3.19 and +3.50. In an interaction model, discipline-specific AI effects were +11.5 for dietetics (94% ETI +6.8 to +16.3), +2.7 for physiotherapy (-1.9 to +7.4), +0.9 for occupational therapy (-3.7 to +5.6) and -1.3 for speech and language therapy (-5.7 to +3.2). These exploratory estimates agree with the descriptive discipline pattern and show that the pooled effect was driven principally by dietetics.

### Table S21. Nesting of evaluators and note writers

| Factor | Levels | Levels per discipline | Levels contributing to more than one discipline |
|---|---|---|---|
| Evaluator | 8 | 2 / 2 / 2 / 2 | 0 |
| Note writer | 8 | 2 / 2 / 2 / 2 | 0 |

Evaluators and note writers were fully nested within discipline. Within each discipline, the same two professionals wrote and evaluated notes reciprocally, so the evaluator and writer effects were largely aliased. Their separate posterior SDs changed with the prior, whereas the note-type estimate did not. The combined professional-level SD was 7.4 (94% ETI 4.1 to 15.7). Because both professional factors were nested within discipline, the combined term also absorbed between-discipline differences. Adding discipline as a fixed effect reduced the separate variance components but did not materially change the note-type estimate.

### Table S22. Sample-size sensitivity and planning values

| Quantity | Value |
|---|---|
| Observed paired difference (SD) | +3.40 (7.24) |
| Achieved 95% CI half-width | +/-3.39 points |
| Minimum detectable dz at n=20, 80% / 90% power | 0.66 / 0.76 (4.8 / 5.5 points) |
| Pairs needed for dz=0.47, 80% / 90% power | 38 / 50 |
| Pairs needed for dz=0.55, 80% / 90% power | 28 / 37 |
| Pairs needed for dz=0.30, 80% / 90% power | 90 / 119 |
| Bootstrap Wilcoxon power at n=10 / 20 / 30 / 40 / 60 | 0.25 / 0.53 / 0.74 / 0.86 / 0.96 |

No a priori sample-size calculation was performed; the sample was determined by feasibility. The planning values are provided for future discipline-specific studies. The bootstrap calculation resampled observed paired differences after re-centring on the observed mean and should be interpreted as a planning aid rather than post hoc power for this study.

### Table S23. Error densities with 95% paired-bootstrap confidence intervals

| Outcome | AI | Professional | Difference (professional - AI), 95% CI | Ratio (professional:AI), 95% CI |
|---|---|---|---|---|
| All errors | 3.35 | 8.55 | 5.20 (3.85 to 6.55) | 2.55 (1.93 to 3.59) |
| Omissions | 1.45 | 5.20 | 3.75 (2.80 to 4.65) | 3.59 (2.47 to 6.06) |
| Critical+High | 0.35 | 0.75 | 0.40 (-0.10 to 1.00) | 2.14 (0.67 to 4.60) |

Confidence intervals were estimated by resampling paired consultations 10,000 times. At least one Critical+High error occurred in seven AI notes and six professional notes. The professional note contained more severe errors in six consultations, the AI note in four and 10 were tied (paired Wilcoxon p=0.22). The professional arm's higher density arose from clustering of several severe errors within a small number of notes. The all-error and omission intervals excluded no difference; the Critical+High interval did not. At the observed serious-error contrast, paired-comparison power was approximately 18%, and approximately 120 pairs would be required for 90% power under bootstrap resampling of the observed consultation pairs.

#### *Evaluation pipeline and severity rubric*

#### Table S24. Evaluation pipeline and model configuration

| Stage | Model | Configuration and decision rule |
|---|---|---|
| Automated reviewer | GPT-5.5 | Bidirectional note-to-transcript and transcript-to-note mapping, followed by entity-level validation; pinned internal evaluation-tool version |
| Professional flags to atomic claims | GPT-5.5 | A deterministic pre-filter normalised responses indicating that no issue was found |
| Adjudication of professional claims | GPT-5.5 | Required a verbatim transcript span; ambiguous evidence defaulted in the professional's favour |
| False-positive screen: unsupported statements | GPT-5.5 | Genuine, paraphrase-supported, reasonable inference, benign detail, terminology variant or uncertain |
| False-positive screen: omissions | GPT-5.5 | Genuine, present/reworded, not clinically significant, appropriate concision or uncertain |
| Targeted severe-error screen | GPT-5.5 | Applied to all 22 Critical+High flags |
| Severity panel | GPT-5.5 and Claude Opus 4.8; Gemini 2.5 Pro excluded post hoc | Batches of 25 errors; each error required a grade from every declared panel member |

All evaluation stages were blinded to note authorship, and the transcript reference excluded the non-spoken scenario header. Each rater scored severity and likelihood from 1 to 5 in context. Severity represented the potential clinical consequence if believed: 1 negligible, 2 minor, 3 moderate, 4 major and 5 catastrophic. Likelihood represented the chance that the error would survive review and propagate to harm: 1 very unlikely, 2 unlikely, 3 possible, 4 likely and 5 very likely. The product defined Low (1-4), Moderate (5-9), High (10-15) and Critical (16-25). If the two declared raters disagreed on tier, the more severe tier was retained. The rubric directed raters to score up negation or status flips, medication, dose, unit or laterality changes, diagnosis substitutions and wrong identifiers, while scoring formatting variants and obvious benign inferences lower. GPT-5.5 was used in the automated reviewer, false-positive screening and the declared severity panel; these stages therefore were not independent. Verbatim prompts are withheld because they form part of proprietary study-analysis and evaluation tooling; requests may be directed to the corresponding author.

Evaluators completed an approximately 10-minute training video on the PDQI-9 and adjudication process before the session; note writers completed a separate approximately 10-minute training video. The videos may be requested from the corresponding author subject to commercial and governance review.

All participants provided written informed consent to participate, for research use of the resulting notes and recordings, and for publication of the de-identified excerpts in Tables S6 and S9. The scripted encounters were wholly synthetic and did not derive from real patient encounters, patient records or patient data.

### Supplementary figure S1

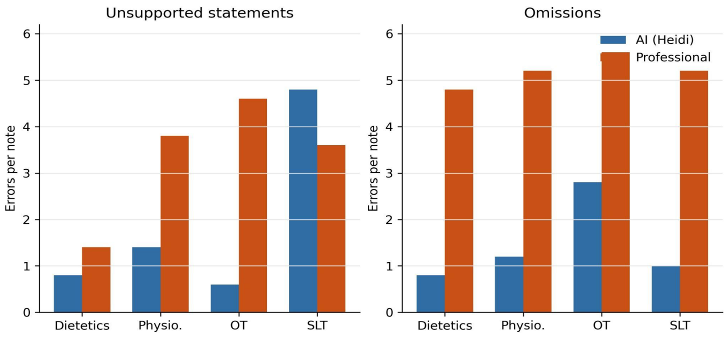


Figure S1. Unsupported-statement and omission density by discipline and note author in the automated-reviewer arm. Physio., physiotherapy; OT, occupational therapy; SLT, speech and language therapy. Values correspond to Supplementary Table S7.
