## Supplementary material for "Quality and transcript-grounded clinical-risk error burden of AI-generated vs allied health professional clinical notes: a paired simulation study across four disciplines": DECIDE-AI Checklist

Bergman H, Liu V, Ali S *Page references: M = main manuscript; S = supplementary material.*

#### Title and abstract

| Item | Theme | DECIDE-AI recommendation | Page |
|---|---|---|---|
| 1 | Title | Identify the study as early clinical evaluation of a decision support system based on AI or machine learning, specifying the problem addressed. | M1 |
| I | Abstract | Provide a structured summary of the study. Consider including: intended use of the AI system, type of underlying algorithm, study setting, number of patients and users included, primary and secondary outcomes, key safety endpoints, human factors evaluated, main results, conclusions. | M2 |

#### Introduction

| Item | Theme | DECIDE-AI recommendation | Page |
|---|---|---|---|
| 2 | Intended use | a) Describe the targeted medical condition(s) and problem(s), including the current standard practice, and the intended patient population(s). b) Describe the intended users of the AI system, its planned integration in the care pathway, and the potential impact, including patient outcomes, it is intended to have. | M3-4, 14 |
| II | Objectives | State the study objectives. | M2-M3 |

#### Methods

| Item | Theme | DECIDE-AI recommendation | Page |
|---|---|---|---|
| III | Research governance | Provide a reference to any study protocol, study registration number, and ethics approval. | M5-7; S16 |
| 3 | Participants | a) Describe how patients were recruited, stating the inclusion and exclusion criteria at both patient and data level, and how the number of recruited patients was decided. b) Describe how users were recruited, stating the inclusion and exclusion criteria, and how the intended number of recruited users was decided. c) Describe steps taken to familiarize the users with the AI system, including any training received prior to the study. | a) N/A; b) M4, 6; c) M4; S20 |
| 4 | AI system | a) Briefly describe the AI system, specifying its version and type of underlying algorithm used. Describe, or provide a direct reference to, the characteristics of the patient population on which the algorithm was trained and its performance in preclinical development/validation studies. b) Identify the data used as inputs. Describe how the data were acquired, the process needed to enter the input data, the pre-processing applied, and how missing/low-quality data were handled. c) Describe the AI system outputs and how they were presented to the users (an image may be useful). | a) M4, 15; S20; b) M4-5; S20; c) M4 |
| 5 | Implementation | a) Describe the settings in which the AI system was evaluated. b) Describe the clinical workflow/care pathway in which the AI system was evaluated, the timing of its use, and how the final supported decision was reached and by whom. | a) M4; b) M4, 14 |
| IV | Outcomes | Specify the primary and secondary outcomes measured. | M5 |

| | | | |
|---|---|---|---|
| 6 | Safety and errors | a) Provide a description of how significant errors/malfunctions were defined and identified. b) Describe how any risks to patient safety or instances of harm were identified, analyzed, and minimized. | a) M4-5; b) M5-6, 9-15; S3-15 |
| 7 | Human factors | Describe the human factors tools, methods or frameworks used, the use cases considered, and the users involved. | N/A; M4, 15 |
| V | Analysis | Describe the statistical methods by which the primary and secondary outcomes were analyzed, as well as any pre-specified additional analyses, including subgroup analyses and their rationale. | M5-6; S16-20 |
| 8 | Ethics | Describe whether specific methodologies were utilized to fulfil an ethics-related goal (such as algorithmic fairness) and their rationale. | N/A |
| VI | Patient involvement | State how patients were involved in any aspect of: the development of the research question, the study design, and the conduct of the study. | M7 |

### Results

| Item | Theme | DECIDE-AI recommendation | Page |
|---|---|---|---|
| 9 | Participants | a) Describe the baseline characteristics of the patients included in the study, and report on input data missingness. b) Describe the baseline characteristics of the users included in the study. | a) N/A; missingness M6; b) M4; S18-19 |
| 10 | Implementation | a) Report on the user exposure to the AI system, on the number of instances the AI system was used, and on the users' adherence to the intended implementation. b) Report any significant changes to the clinical workflow or care pathway caused by the AI system. | a) M4; b) N/A |
| VII | Main results | Report on the pre-specified outcomes, including outcomes for any comparison group if applicable. | M7-11; S2-20 |
| VIII | Subgroups analysis | Report on the differences in the main outcomes according to the pre-specified subgroups. | M7-10; S2-3, 9-10 |
| 11 | Modifications | Report any changes made to the AI system or its hardware platform during the study. Report the timing of these modifications, the rationale for each, and any changes in outcomes observed after each of them. | M6, 15; S16, 20 |
| 12 | Human-computer agreement | Report on the user agreement with the AI system. Describe any instances of and reasons for user variation from the AI system's recommendations and, if applicable, users changing their mind based on the AI system's recommendations. | N/A; M4, 15 |
| 13 | Safety and errors | a) List any significant errors/malfunctions related to: AI system recommendations, supporting software/hardware, or users. Include details of: (i) rate of occurrence, (ii) apparent causes, (iii) whether they could be corrected, and (iv) any significant potential impacts on patient care. b) Report on any risks to patient safety or observed instances of harm (including indirect harm) identified during the study. | M9-15; S3-15 |
| 14 | Human factors | a) Report on the usability evaluation, according to recognized standards or frameworks. b) Report on the user learning curves evaluation. | N/A; M4, 15 |

### Discussion

| Item | Theme | DECIDE-AI recommendation | Page |
|---|---|---|---|
| 15 | Support for intended use | Discuss whether the results obtained support the intended use of the AI system in clinical settings. | M14-15 |

| | | | |
|---|---|---|---|
| 16 | Safety and errors | Discuss what the results indicate about the safety profile of the AI system. Discuss any observed errors/malfunctions and instances of harm, their implications for patient care, and whether/how they can be mitigated. | M9-15 |
| IX | Strengths and limitations | Discuss the strengths and limitations of the study. | M15 |

### Statements

| Item | Theme | DECIDE-AI recommendation | Page |
|---|---|---|---|
| 17 | Data availability | Disclose if and how data and relevant code are available. | M16 |
| X | Conflicts of interest | Disclose any relevant conflicts of interest, including the source of funding for the study, the role of funders, any other roles played by commercial companies, and personal conflicts of interest for each author. | M16 |

### Reference

Vasey B, Nagendran M, Campbell B, et al. Reporting guideline for the early-stage clinical evaluation of decision support systems driven by artificial intelligence: DECIDE-AI. BMJ. 2022;377:e070904. doi:10.1136/bmj-2022-070904. Published concurrently as Nat Med. 2022;28(5):924-933. doi:10.1038/s41591-022-01772-9. Item wording in this checklist is quoted from the DECIDE-AI reporting item checklist supplied with the Nature Medicine article.
